# Immune Heterogeneity and Epistasis Explain Punctuated Evolution of SARS-CoV-2

**DOI:** 10.1101/2022.07.27.22278129

**Authors:** Bjarke Frost Nielsen, Yimei Li, Kim Sneppen, Lone Simonsen, Cécile Viboud, Simon A. Levin, Bryan T. Grenfell

## Abstract

Identifying drivers of viral diversity is key to understanding the evolutionary as well as epidemiological dynamics of the COVID-19 pandemic. Using rich viral genomic data sets, we show that periods of steadily rising diversity have been punctuated by sudden, enormous increases followed by similarly abrupt collapses of diversity. We introduce a mechanistic model of saltational evolution with epistasis and demonstrate that these features parsimoniously account for the observed temporal dynamics of inter-genomic diversity. Our results provide support for recent proposals that saltational evolution may be a signature feature of SARS-CoV-2, allowing the pathogen to more readily evolve highly transmissible variants. These findings lend theoretical support to a heightened awareness of biological contexts where increased diversification may occur. They also underline the power of pathogen genomics and other surveillance streams in clarifying the phylodynamics of emerging and endemic infections. In public health terms, our results further underline the importance of equitable distribution of up-to-date vaccines.

## Introduction

During the coronavirus disease 2019 (COVID-19) pandemic, the responsible pathogen, severe acute respiratory syndrome coronavirus 2 (SARS-CoV-2), has continuously evolved. However, evolution has by no means happened at an even pace, but rather through a pattern of steady diversification punctuated by sudden large jumps involving dozens of point mutations. Indeed, it has been suggested that SARS-CoV-2 exhibits saltational evolution, a process where evolution proceeds by large multimutational jumps, rather than gradually (***Corey et al., 2021***).

A simple way to quantify the genomic diversity existing at a given time is through the pairwise Hamming distance. Given two genomes, the pairwise Hamming distance simply measures how many nucleotides the two sequences disagree on. This rather crude measure turns out to reveal surprisingly robust patterns of viral diversification.

Due to the large amount of full genome sequencing performed on SARS-CoV-2 specimens during the COVID-19 pandemic, Hamming distances can be computed not just at the level of summary statistics, but as temporally varying distributions (Figure 1; Video 1), revealing a pattern of slowly increasing diversity punctuated by abrupt increases and subsequent collapses in diversity.

**Figure 1.**
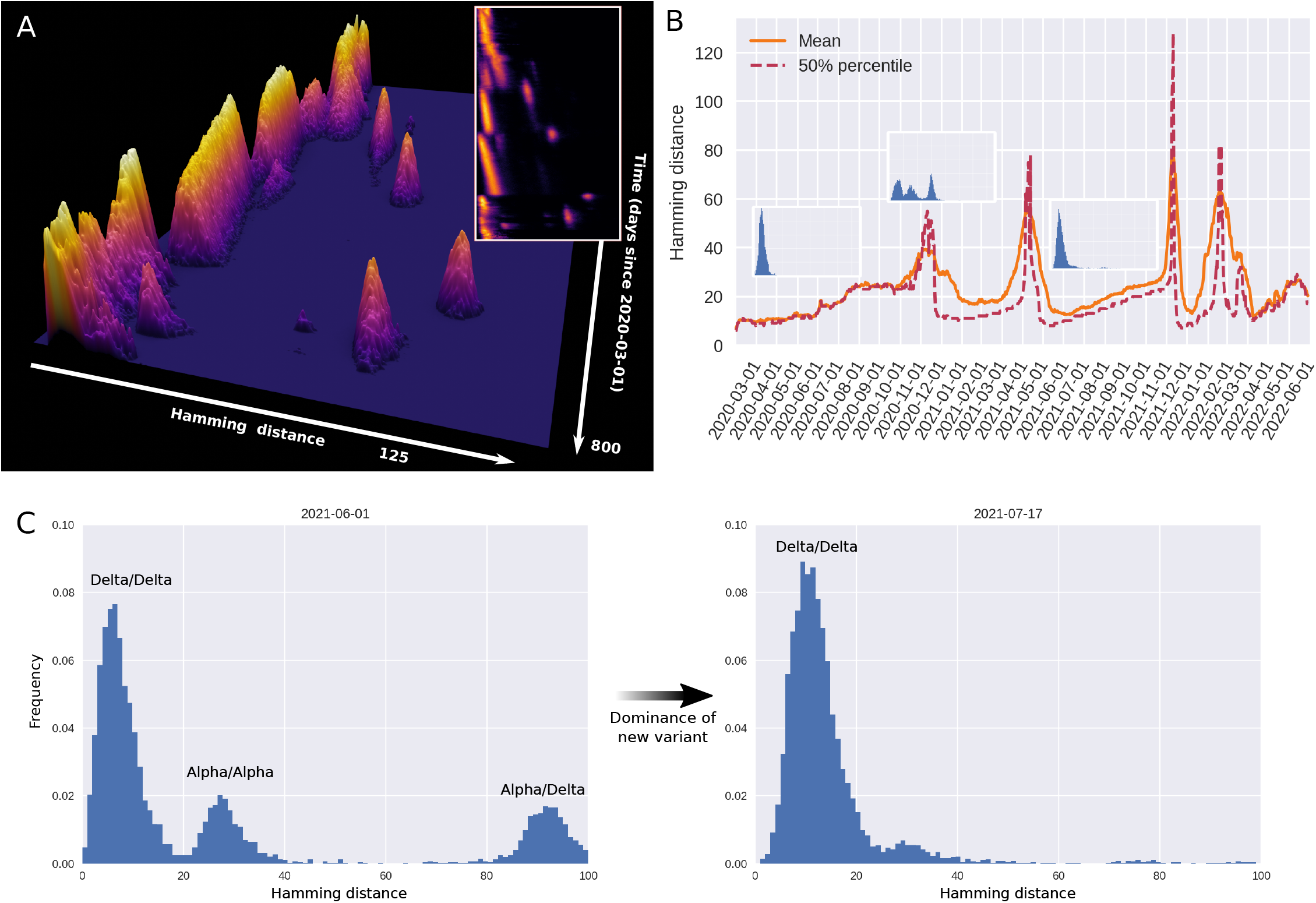
Genomic diversity over time in SARS-CoV-2, UK genomic sureveillance data. **(A)** Full, time-dependent Hamming distance distribution between 2020-03-01 and 2022-05-10 (UK data, GenBank via Nextstrain (***Hadfield et al., 2018***)). Insert: Two-dimensional heatmap representation of the time-dependent Hamming distribution. **(B)** Time evolution of the mean and median Hamming distance. Each time point represents Hamming distances between genomes sampled within a one-week window beginning on that date. The three miniature inserts show Hamming distance histograms at three different time points. **C)** Left: A snapshot of the Hamming distance distribution for genomes sampled during a one-week window starting on June 1st, 2021. The three distinct peaks correspond to the Hamming distances between pairs of genomes from each of the prevailing variants at the time, Alpha and Delta. Right: 46 days later, a single variant (Delta) dominates. See also **Video 1** for the animated Hamming histogram. **Figure 1–Figure supplement 1**. Data analysis workflow. **Figure 1–Figure supplement 2**. Hamming distributions for influenza H3N2 and H1N1.

That much can be gleaned from considering the time-development of the mean (or median) Hamming distance. However, the dynamics of the often multimodal distribution is not captured by the mean Hamming distance, even if temporally resolved, and much less by the usual static treatment. The full, time-dependent Hamming distribution possesses further structure that reveals that successive variants are well-separated in sequence space, suggesting that one did not arise from the other by a string of single-point mutations accruing in successive hosts. Rather, an evolutionary jump – a saltation – seems to have taken place at each major transition (see Video 1).

### Dynamical explanations

There are several plausible mechanisms that may contribute to saltational evolution in SARS-CoV-2, including increased build-up of mutations in immunocompromised individuals infected with SARS-CoV-2 (***Corey et al., 2021***; ***Nussenblatt et al., 2022***; ***Avanzato et al., 2020***; ***Choi et al., 2020***; ***Truong et al., 2021***; ***Kemp et al., 2021***; ***Harari et al., 2022***; ***Kumata and Sasaki, 2022***) and evolution in animal reservoirs followed by animal-to-human transmission (***Kupferschmidt, 2021***; ***Larsen et al., 2021***).

In this paper, we present a mathematical model aimed at capturing the particular punctuated evolutionary pattern of SARS-CoV-2. Our goal is to recapitulate the main features of the temporal Hamming distribution observed during the COVID-19 pandemic (see Figure 1A) as parsimoniously as possible in a dynamical model.

We show that the overall pattern can be captured by combining epistasis with heterogeneous within-host evolution. The model is sufficiently general that it does not make any assumptions about the biological mechanism behind saltations.

The proposed model is conceptually related to the NK Model of ***Kauffman and Levin*** (***1987***) and ***Kauffman and Weinberger*** (***1989***) in that it operates on the space of possible genotypes, with each genotype corresponding to a preassigned fitness value. This is in contrast to phenotypic fitness landscape models which operate directly on the space of possible values of some quantifiable trait. The most well-known among those is perhaps Fisher’s geometric model (***Fisher, 1930***) which assumes a continuous phenotypic (‘trait’) space with a single optimum (***Blanquart and Bataillon, 2016***) and that the effects of single mutations are mild (***Martin, 2014***). The NK Model, a genotypic fitness landscape model, instead explicitly allows for a rough (epistatic) fitness landscape. The NK model, however, does not include the concept of *neutral space* – in that model, mutations are generically accompanied by a change in fitness. Our model includes neutral mutations and is, in that respect, closer to the models of ***Koelle et al***. (***2006***); ***Newman and Engelhardt*** (***1998***).

However, a crucial component of our model is the presence of sign epistasis, i.e. that the fitness contribution of a point mutation may change sign (going from deleterious to beneficial or viceversa) depending on the presence of other mutations. This property turns out to offer an explanation for the role of saltation in evolving high-fitness SARS-CoV-2 genotypes.

In a recent study, ***Starr et al***. (***2022***) showed by deep mutational scanning that epistasis – including sign epistasis – is an important important feature of SARS-CoV-2 evolution. As a concrete example, they show that the N501Y mutation (which is present in the Alpha, Beta and Omicron variants) and Q498R exhibit sign epistasis. In this case, the presence of the N501Y substitution changes the contribution of Q498R from deleterious to advantageous, as measured by angiotensin-converting enzyme 2 (ACE2) receptor binding affinity.

In general, the fitness landscape of an organism is combinatorially large, and the number of possible evolutionary paths from one genotype to another fitter one is, a priori, enormous. However, in a seminal paper, ***Weinreich et al***. (***2006***, 2013) showed that only very few such paths are in fact accessible. The interpretation of this finding in terms of fitness landscapes is that epistasis or the *ruggedness* of the landscape is highly important for understanding evolutionary trajectories (***Blanquart and Bataillon, 2016***). However, even if evolutionary paths seem blocked, this conclusion may only hold in the weak mutation limit, i.e. when the probability of multiple mutations arising in the same genome within a generation is low (***Katsnelson et al., 2019***). If saltational evolution is possible, even seemingly inaccessible regions of the fitness landscape may be explored by the organism. Our model suggests that such saltations may thus increase – or, in some cases, altogether enable – the emergence of new concerning variants.

## Results

### SARS-CoV-2 genomic diversity is characterized by punctuated evolution

On the basis of UK sequences (a particularly rich data set), we have computed a time-dependent Hamming distribution for SARS-CoV-2, which is presented in Figure 1. Figure 1A shows the full Hamming distance histogram as a function of time, from March 2020 to mid-2022, with the colour and height indicating the frequency of observing genome pairs with a particular Hamming distance. The peaks corresponding to saltational variant transitions are clearly visible as isolated ‘islands’ at large Hamming distance. The insert in the same panel shows a 2D representation of the data.

In panel B, time series of the mean and median Hamming distances are shown, revealing clear spikes associated with each of the major variant transitions to date. Each transition event is marked by a very sudden spike in the typical Hamming distance, as is especially clear when considering the median (dashed line). It should be noted that data quality is highest after the end of 2020, when sequencing capacity was greatly increased, and before February 2022. As a concrete example, 4,945 sequences were included for June of 2020, while 72,292 sequences were included for the month of June, 2021.

In Figure 1C (left), a snapshot from June 1st 2021 shows three well-defined peaks. Each peak corresponds to comparisons between pairs of genomes, with the members of each pair belonging to either the Alpha or Delta variant. The peak corresponding to the highest Hamming distance is of course that due to comparisons between the ‘new’ and ‘old’ variant, since these are furthest from each other in a genomic sense. Similarly, the plot clearly shows that variation within the Delta variant is, at that point in time, much lower than within the Alpha variant, since each of the Delta variant genomes belong to a clade with a recent common ancestor. In the right half of Figure 1C, the situation 46 days later is shown, once the Hamming distribution has collapsed to a single peak, corresponding to the then-dominant Delta variant.

During the month of March, 2020, the Hamming distribution appears bimodal, but there are no signs of saltation. This transient bimodality, present in the early pandemic, can most clearly be seen in Video 1. This can be explained by the D614G substitution, which was associated with a clade that dominated from around the end of March/beginning of April 2020 (***Volz et al., 2021***). This early, saltation-free transition is reminiscent of a result by ***Kauffman and Levin*** (***1987***), who suggested that adaptation on a rugged fitness landscape is associated with two separate time scales. First, the pathogen searches its neighbourhood in the fitness landscape until it finds a local maximum. This does not require saltation and happens rather rapidly. Then, on a slower time scale, the pathogen may transition to new fitness peaks by saltation.

Due to the relatively high quality of SARS-CoV-2 genomic surveillance in the United Kingdom, both in terms of the absolute number of publicly available sequences and per capita coverage, we have based the bulk of our observations on UK sequence data. However, patterns similar to those presented here can be observed in US data, the analysis of which is included in Appendix 1.

For each day in the included range (2020-03-01 to 2022-05-10) a 7-day window (consisting of the indicated day and the 6 following days) was considered. All high-quality sequences obtained within that window were pooled, and a distribution of Hamming distances was compiled by repeatedly picking out random pairs from the sequence pool and comparing.

While the Hamming distance is a somewhat crude measure of the variance between circulating genomes, it turns out to offer a surprisingly powerful window into the evolution of SARS-CoV-2 when large amounts of sequence data are available. The transitions ancestral^1^ variant→Alpha, Alpha→Delta, Delta→Omicron (BA.1) as well as Omicron BA.1→BA.2 all show the tell-tale signs of saltational evolution. This is perhaps especially clear when considering the median Hamming distance (Figure 1B, dashed line) which increases almost discontinuously at these transitions, while the full distribution (Figure 1A) shows clearly defined, disconnected ‘islands’. The Omicron BA.2→BA.5 transition is somewhat less clearly defined, although a moderately sized saltation does appear to be present in the data. It should be noted that UK sequencing has become less dense since February 2022, explaining why the picture is somewhat murkier for the BA.2→BA.5 transition^2^.

**Video 1.**
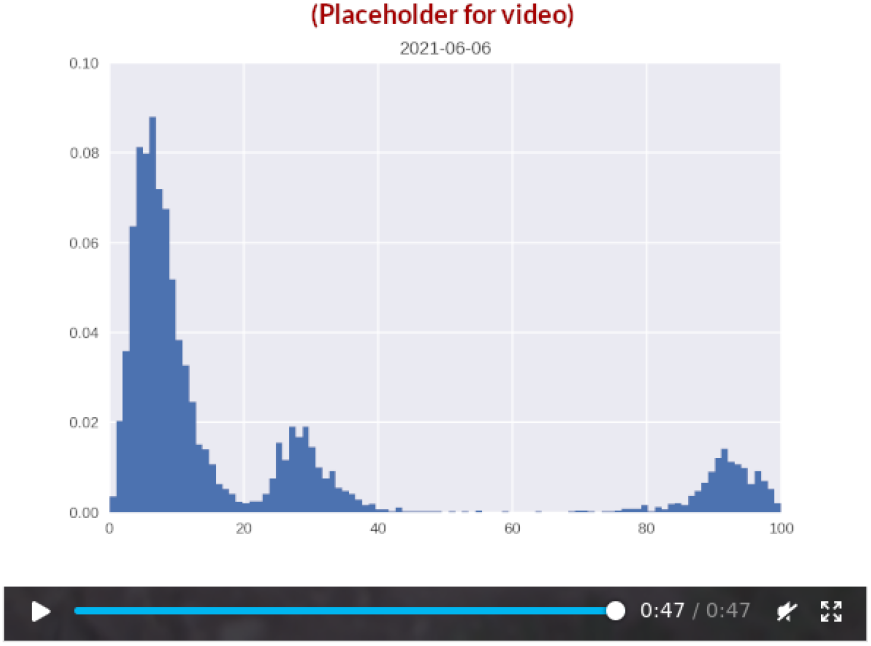
Day-by-day time development of the Hamming distribution for UK samples obtained between March 2020 and May 2022. Each snapshot is based on samples obtained within a one-week time window. **Click image to access video**.

As shown in Appendix 2 Figure 1, all but one of these saltational transitions are also associated with a discontinuous increase in the distance to the *origin* (Wuhan-Hu-1, GenBank reference sequence accession number MN908947.3). The exception is the Alpha→Delta transition, where a moderate decrease is observed. In other words, the Delta variant is closer to the ancestral variant than Alpha is. In Appendix 2, we model one possible explanation for this phenomenon, namely the occurrence of persistent infections.

The plots of Figure 1 are based on the entire SARS-CoV-2 genome, meaning that a substitution leading to an amino acid change in the spike protein (a major antigen) counts just as much as a synonymous mutation elsewhere in the genome. In Figure 2, we probe to what extent the observed drift-boom-bust pattern of diversity is driven by changes in the S-gene (coding for the spike protein) or by (non-)synonymous mutations. Overall, the pattern is present whether considering only the S-gene (Figure 2B), non-synonymous mutations (2C) or the entire genome (2A). We interpret this to mean that

**Figure 2.**
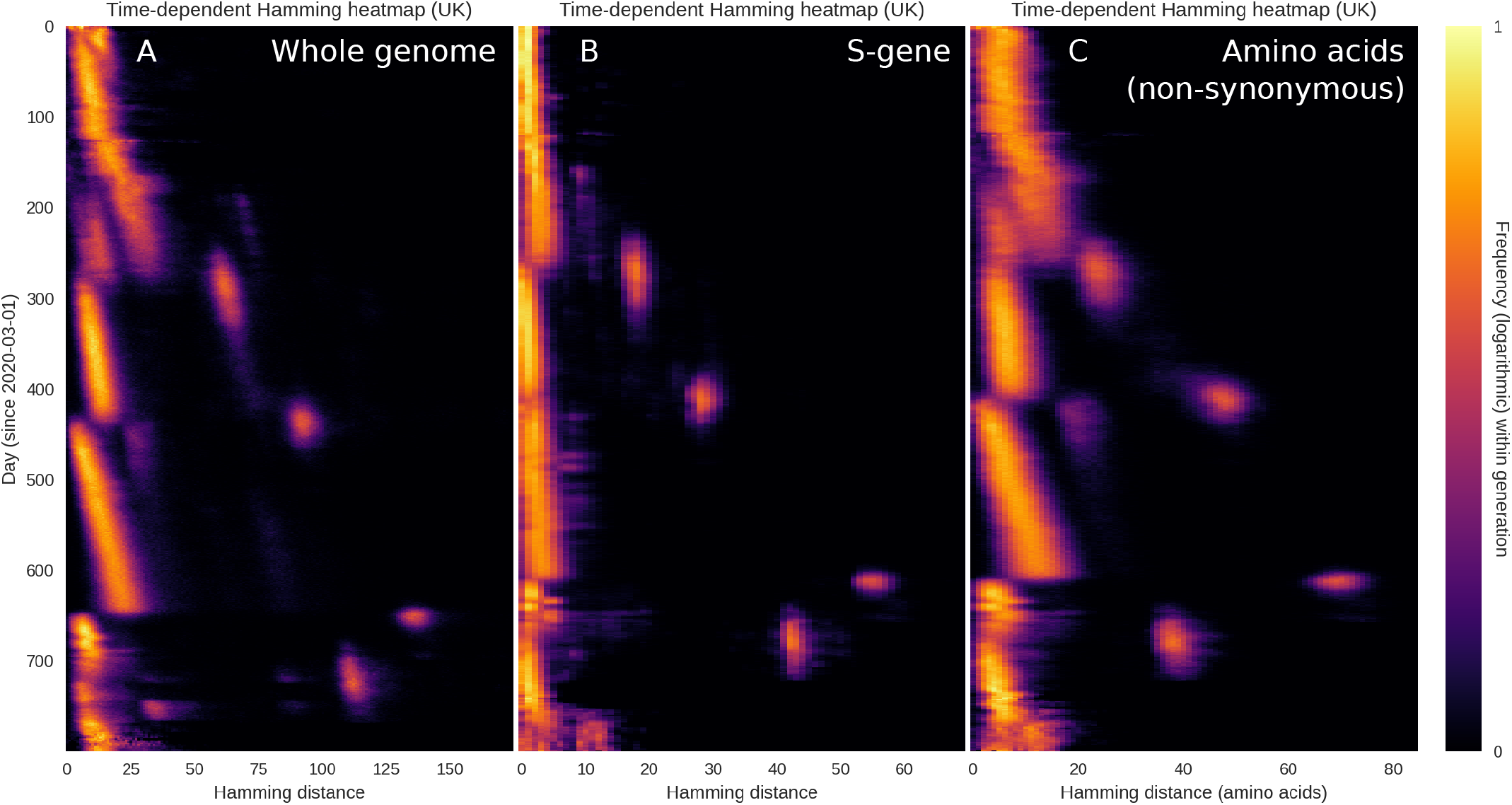
Restricting the Hamming distribution to the S-gene or the amino acid sequence. The overall temporal pattern of diversity seen in Figure 1 is found to persist when the analysis is restricted to the S-gene or non-synonymous mutations. **(A)** Temporal Hamming distance distribution based on the whole genome, included for reference. For each time on the vertical axis, the colour encodes the histogram of Hamming distances between genomes sampled within a one-week window starting on that date. **(B)** Temporal Hamming distance distribution confined to the S-gene which encodes the SARS-CoV-2 Spike protein. **(C)** Time evolution of the Hamming distance distribution as measured by the number of amino acid changes. We use this as a proxy for non-synonymous mutations, since a synonymous mutation would not produce an amino acid change.

1. The drift seen between saltations is not driven solely by synonymous mutations but affects the amino acid sequence as well.
2. When saltations occur, mutations are observed within the spike protein as well as outside it.
3. The observed pattern is quite robust, being observed within the whole genome, in the amino acid sequence as well as within the S-gene itself.

It is notable, however, that the S-gene does not undergo quite as much drift as the whole genome, relatively speaking. That is to say, when the whole genome is considered, the Delta→Omicron jump is associated with a peak that is approximately 5.5 times larger than the typical Hamming distances in the weeks that preceded it, while the ratio is closer to 11 for the spike protein. We interpret this to mean that, while the S-gene is subject to large saltations, it undergoes less drift than an average, similarly sized section of the genome.

### Mechanistic modelling captures the essential dynamics

Our goal is to capture the overall temporal pattern of diversity observed in Figure 1 in a mathematical model that is as parsimonious as possible. The model we have formulated consists of two parts: a branching process and an evolutionary algorithm incorporating sign epistasis and saltational evolution. The details of both of these elements can be found in the Methods and Materials section. See also Figure 3–Figure supplement 1 for a schematic description of the model elements.

The model assumes the existence of a number of possible high-fitness genotypes, but that each of them are ‘screened’ by epistasis. From a fitness landscape viewpoint, this can be thought of as a landscape with a number of peaks, each of which is surrounded by a fitness trough or valley. The extent of sign epistasis is then determined by the depth (and width) of these valleys.

To get from a moderate-fitness genotype to a local fitness peak, it is thus necessary to either traverse a region of low fitness, with its potential for extinction, or to somehow jump across that valley.

Evolutionary models typically assume that the ‘weak mutation limit’ holds, meaning that the probability of several mutations arising in one genome in one generation is negligible (***Katsnelson et al., 2019***), leading to gradual evolution. However, as described in the introduction, there are several mechanisms which can introduce a sudden burst of novelty within a single host, including by recombination (***Burel et al., 2022***; ***Duerr et al., 2022***; ***Jackson et al., 2021***). The most well-documented is perhaps elevated mutation in immunocompromised individuals. Our model, however, is agnostic with respect to the precise etiology, but includes saltation simply as rare occurrences of drastically increased evolution within a single host.

As shown in Figure 3, the model replicates the main features observed in Figure 1, including the long periods of drift (linearly increasing pairwise Hamming distances) punctuated by rapid rises and subsequent collapses of diversity. Just as in the empirical data, each variant transition is accompanied by three distinct peaks in the Hamming distribution.

**Figure 3.**
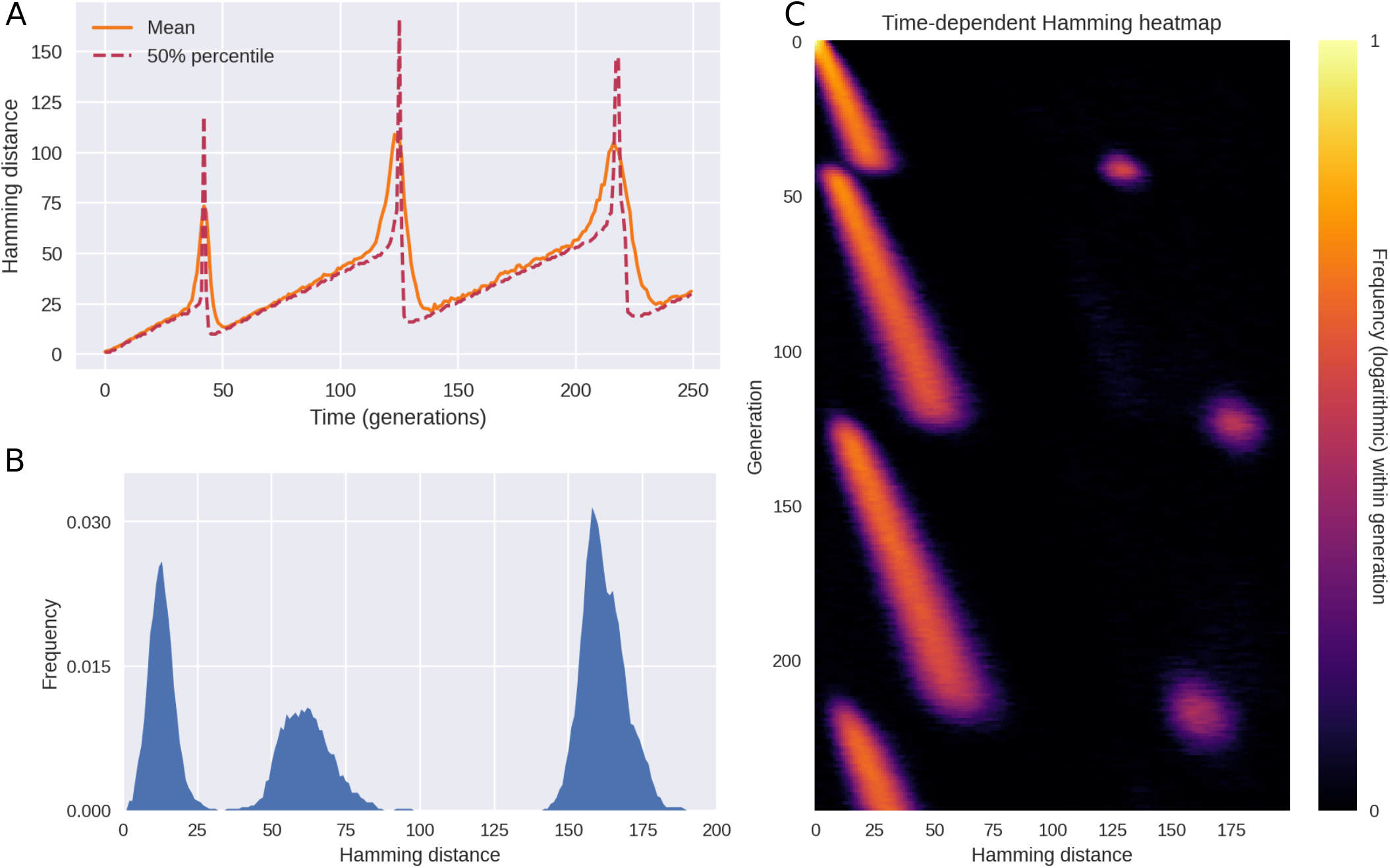
Simulated outbreak with saltation (heterogeneous mutation rates) and epistasis. **(A)** Time evolution of the mean and median Hamming distance between bitstring genomes present in any given generation of the model simulation. The pattern of genetic drift punctuated by sudden increases and subsequent collapses in diversity is similar to what is observed in SARS-CoV-2 (see Figure 1). **(B)** A snapshot of the Hamming distance distribution in generation *t* = 218 of the simulated outbreak. Just as in Figure 1, the three distinct peaks correspond to the distances between pairs of genomes from each of the two prevailing variants at the time. **(C)** Time evolution of the full Hamming distance distribution. For each generation on the vertical axis, the colour encodes the histogram of Hamming distances between genomes within that generation. The parameters used in these simulations were *ϵ* = 0.001, *d*_0_ = 3, *δR*_*H*_ = 1.0, *δR*_*L*_ = −∞ (i.e. deleterious mutations were fatal to the pathogen). **Figure 3–Figure supplement 1**. Model schematics. **Figure 3–Figure supplement 2**. Coexistence of equally fit variants.

The pattern shown in Figure 3 is the typical outcome of a model simulation, but occasional coexistence of two variants does occur in the model, see Figure 3–Figure supplement 2. This happens when two distinct variants with the same fitness happen to arise close to each other in time.

In the interest of simplicity, we have assumed an epidemic of constant size (constant incidence^3^), however we explore the consequences of relaxing this assumption in the section *Epidemic dynamics and spatial structure*.

If epistasis and saltation are turned off, evolution and variant transitions still happen within the model. The temporal pattern changes, however. In Figure 4, we explore this regime by setting *ϵ*, the frequency of saltations, to zero and letting *δR*_*L*_ = 0, thus disabling sign epistasis. The resulting behaviour is characterized by periods of increasing diversity – essentially, genetic drift – interrupted by sudden collapses of the typical Hamming distance. No sudden spikes are seen in Figure 4A, rendering the dynamics fundamentally different from that of Figures 1 and 3. The behaviour observed in this regime is more reminiscent of the dynamics observed for H3N2 influenza in ***Koelle et al***. (***2006***). However, one could object that the temporal resolution of the empirical time series shown in ***Koelle et al***. (***2006***) is not sufficiently high to allow one to discriminate between the scenarios of our Figures 3 and 4 – after all, the periods of drastically increased pairwise nucleotide Hamming distance seen in Figure 1 are brief and require high temporal resolution to discern. While the amount of genomic data available for SARS-CoV-2 enables this, the picture is murkier for seasonal influenza. In Figure 1–Figure supplement 2, we present the result of applying the analysis of Figure 1A to influenza types H3N2 and H1N1. While there is no apparent evidence of saltation, the available data is relatively coarse-grained.

**Figure 4.**
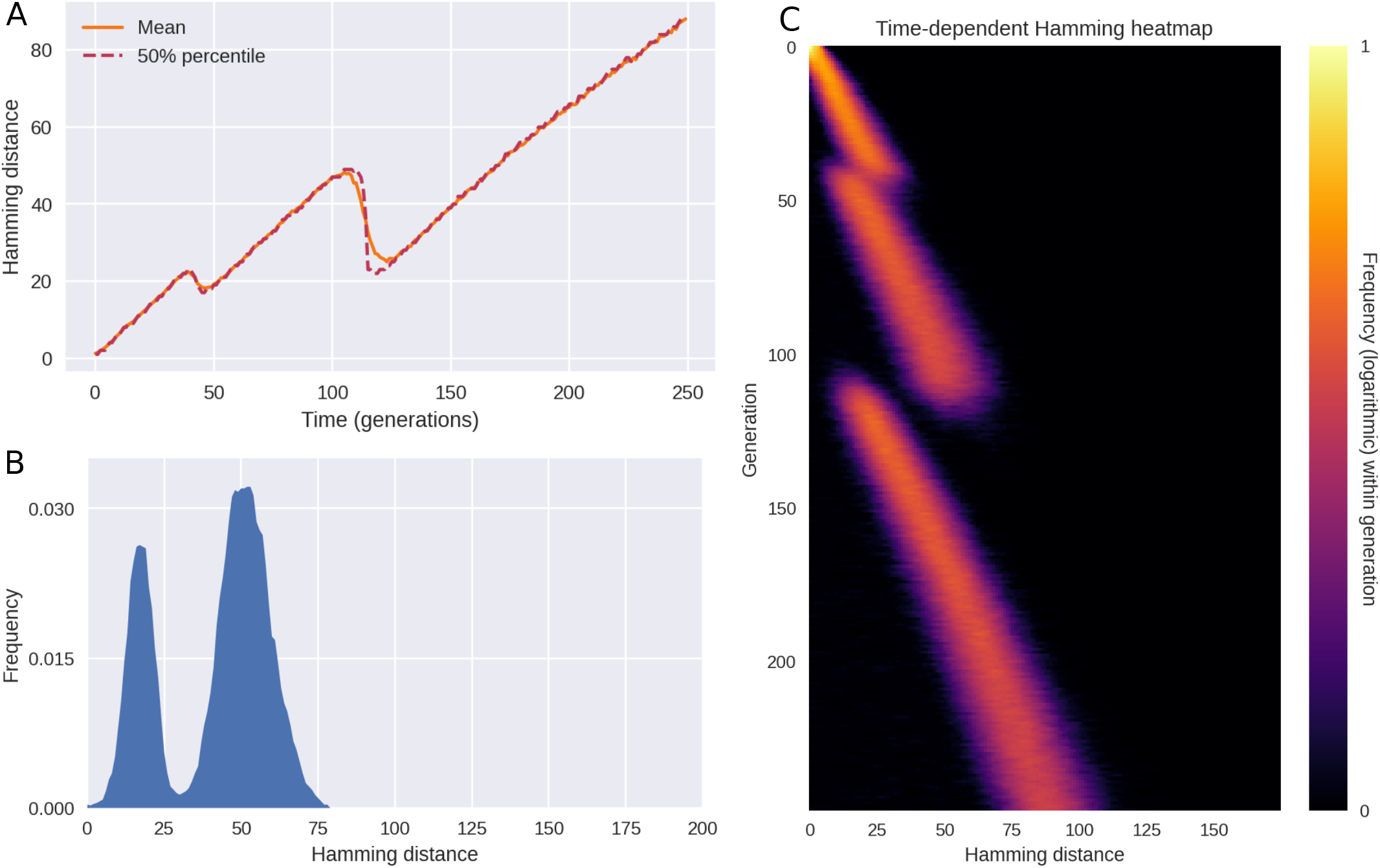
In the absence of epistasis and saltation, model results do not match observations. When saltations do not occur (*ϵ* = 0) and sign epistasis is absent (*δR*_*L*_ = 0), the pathogen will only rarely overcome troughs in the fitness landscape. When it does, the transitions are marked by a collapse of diversity (as measured by the typical Hamming distance), giving a drift-bust-drift dynamics as opposed to the drift-boom-bust pattern seen in SARS-CoV-2. **(A)** Time evolution of the mean and median Hamming distance between genomes present in any given generation of the model simulation. **(B)** A snapshot of the Hamming distance distribution for bitstring genomes at generation *t* = 112 of the simulated outbreak. **(C)** Time evolution of the Hamming distance distribution. For each generation indicated on the vertical axis, the colour encodes the histogram of Hamming distances between genomes within that generation. **Figure 4–Figure supplement 1**. Saltational evolution in the absence of sign epistasis.

Another influential evolutionary model of influenza is due to ***Ferguson et al***. (***2003***). In their model, the appearance of new variants is driven by immune system memory and a non-linear relation between Hamming distance and cross-immunity, the latter in the form of short-lived strain-transcending immunity. While a sensible model for seasonal influenza, it gives rise to diversity dynamics which are closer to Figure 4 than to the pattern observed for SARS-CoV-2.

In the simulations of Figure 4, saltation and epistasis are completely lacking, but in Figure 4– Figure supplement 1, we consider what happens if some saltational evolution *does* occur, without sign epistasis. Qualitatively, the picture most resembles the saltation-free scenario of Figure 4, but occasional Hamming spikes are observed. Overall, this scenario does not conform to the empirical observations in the form of Figure 1. In the next section, we systematically probe how different levels of epistasis and saltation affects the evolution of new, highly transmissible variants.

Our focus is mainly on the dynamics of diversity, and for this reason we have emphasized the distribution of Hamming distances between viral genomes present in the population at the same time. This goes for the empirical observations (Figure 1) as well as our model simulations (Figure 3). However, in Appendix 2, we explore the distributions of Hamming distance relative to the origin (meaning Wuhan-Hu-1, GenBank reference sequence accession number MN908947.3).

### Saltation facilitates the evolution of highly transmissible variants

Saltational evolution may not only be a way to generate vastly different variants, but may indeed be necessary for the virus to evolve highly fit variants at all. In the presence of strong epistasis, gradual evolution towards a high fitness genotype can be blocked (see Figure 3–Figure supplement 1A). Conceptually, such gradual evolution under strong epistasis would correspond to traversing a deep valley in the fitness landscape by a series of small steps before reaching a peak (***Katsnelson et al., 2019***; ***Smith and Ashby, 2022***). However, such a fitness valley indicates the presence of deleterious mutations which impart a high probability of extinction of the lineage in question, preventing the fitness peak from being reached.

In Figure 5, we explore how the strength of epistasis and the size of saltations affect the ability of the pathogen to evolve new, highly transmissible strains. By the *strength* of epistasis, we mean the typical depth of a valley in the fitness landscape, |*δR*_*L*_|, i.e. the loss in reproductive number suffered.

**Figure 5.**
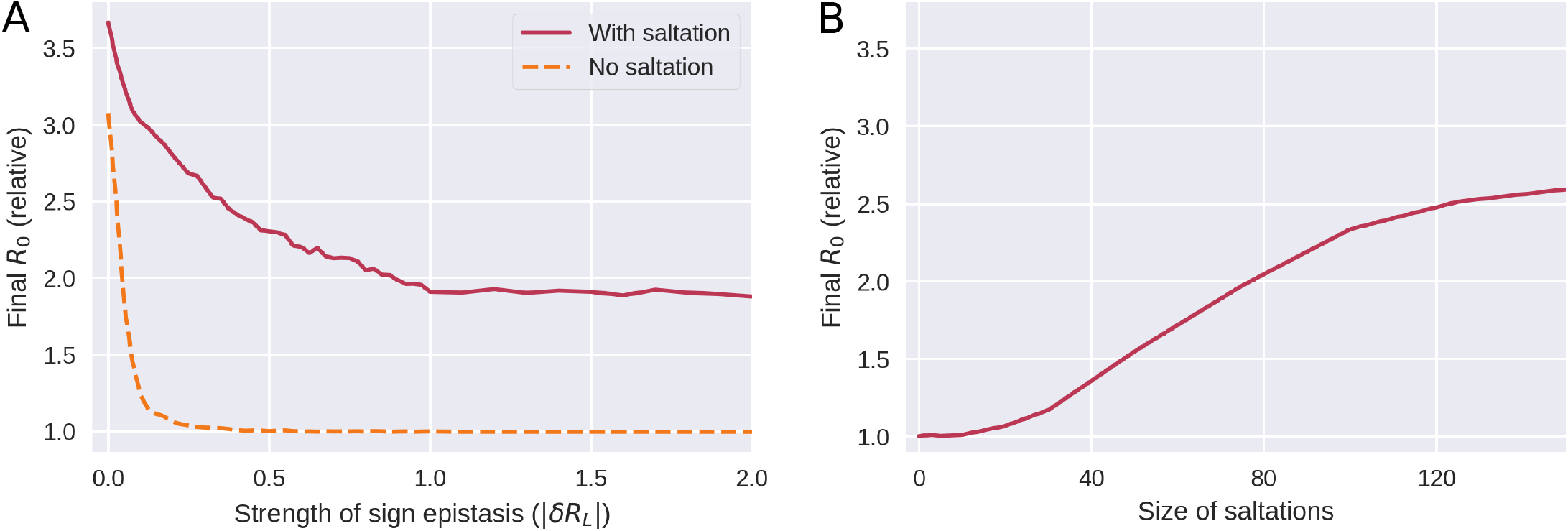
Saltation allows highly transmissible variants to evolve by facilitating evolution across fitness landscape troughs. **A)** Evolution under varying degrees of sign epistasis. The vertical axis indicates the final average reproductive number in the model population after 300 generations of the simulation, relative to (divided by) the reproductive number of the initial variant. The horizontal axis indicates the depth of a valley in the fitness landscape, |*δR*_*L*_|, understood as the reduction in reproductive number suffered due to a deleterious configuration. Here, *δR*_*L*_ was distributed according to a Dirac *δ* distribution and as such its value was deterministic. This panel is based on 90,000 simulations and the parameters used were *d*_0_ = 3 and *δR*_*H*_ = 1. **B)** Evolution with varying degrees of saltation. Moderate sign epistasis is assumed (*δR*_*L*_ = −0.5). All other parameters are as in panel A. This panel is based on 7600 simulations.

For a pathogen which does not undergo saltational evolution (Figure 5A, dashed curve), significant sign epistasis (|*δR*_*L*_| ≳ 0.25 at *d*_0_ = 3 in our simulations) is a roadblock to evolution of high-fitness variants. However, a pathogen which undergoes saltation (fully drawn curve) can overcome this epistatic hindrance. Above a certain threshold (at |*δR*_*L*_| ≈ 1 in Figure 5A), stronger sign epistasis ceases to further impede the emergence of high-fitness variants. The mechanism behind this is that sign epistasis becomes so strong that a fitness valley may be overcome only by pure saltation and is no longer traversable by gradual evolution or a combination of the two.

As shown in Figure 5B, large saltations are necessary to overcome even moderate sign epistasis, further explaining why the Hamming peaks seen in SARS-CoV-2 are so large.

### Epidemic dynamics and spatial structure

In Figure 3, we made a number of simplifying assumptions, the major ones being constant prevalence and absence of any spatial or population structure.

We first relax the former assumption by implementing susceptible-infected-recovered-susceptible (SIRS) dynamics. The infected individuals are now assumed to make up only a fraction of a larger population of total size *N*. Our aim is to ascertain whether the diversity dynamics observed in the previous section are fundamentally altered by allowing a variable number of infected individuals, *I*(*t*), as well as susceptible depletion and waning immunity.

In Figure 6, a typical course of a simulation with SIRS dynamics, epistasis and saltation is shown. As shown in panel A, the number of recovered (immune) individuals varies non-monotonically over time, reflecting that individuals acquire immunity after being infected, and that the immunity eventually wanes. However, as successive variants of greater fitness (greater reproductive number *R*_0_) arise, an endemic plateau is eventually reached. While the epidemiology is very different from that of Figure 3, the Hamming distribution (Figure 6C) is remarkably similar. This indicates that the mechanism of saltational evolution in conjunction with sign epistasis robustly reproduces the punctuated evolutionary dynamics seen in Figure 1.

**Figure 6.**
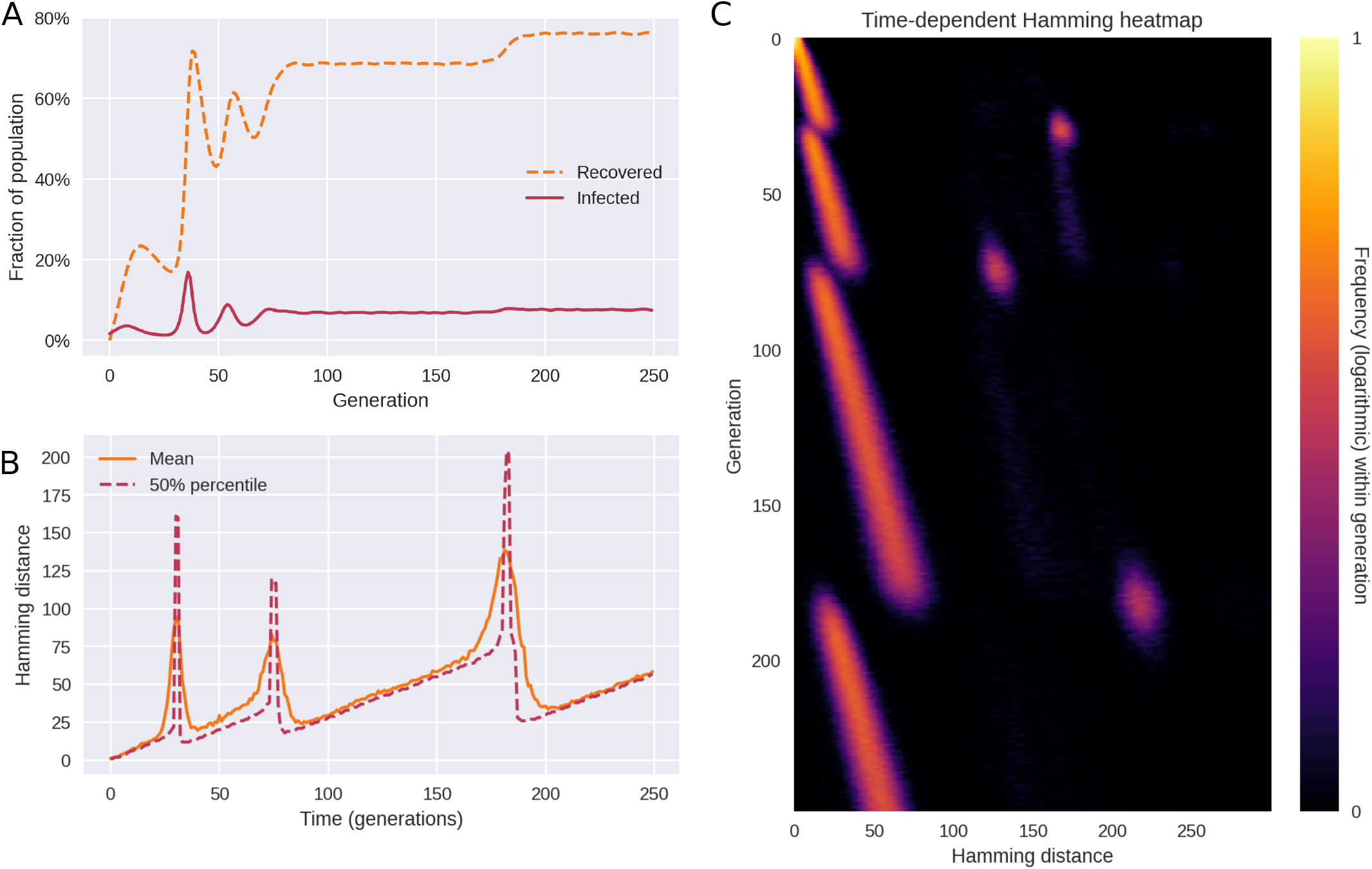
Saltational evolution under susceptible-infected-recovered-susceptible (SIRS) dynamics. The reproductive number of the initial variant is *R*_0_ = 1.2 and immunity wanes at a rate of *ω* = 0.1. Population size is *N* = 6 × 10^5^. The parameters of fitness-altering mutations are *δR*_*H*_ = 1 and *δR*_*L*_ = −∞ (i.e. deleterious mutations are fatal to the organism or prevent transmission). **(A)** Time evolution of the recovered (or immune) fraction of the population. Successive variants have higher reproductive numbers (*R*_0_), eventually leading to two different endemic plateaus. **(B)** Even under variable prevalence, the Hamming dynamics looks similar to that of Figure 3. **(C)** The full temporal Hamming distribution is characterized by the same kind of punctuated evolution as in the simpler constant-prevalence case of Figure 3. **Figure 6–Figure supplement 1**. Simulations with spatial (metapopulation) structure.

In simulations with variable incidence, higher incidence translates to an increased risk of emergence of new variants, all else being equal. Since saltations are simulated as a constant-rate (Poisson) process for each infected individual, the risk of emergence scales with the number of infected. Since simulations are stochastic, this tendency is not necessarily clear from a single realization such as Figure 6. A similar frequency dependence is likely to hold for SARS-CoV-2, since rare occurrences in terms of within-host evolution are proportionally more likely to be observed with higher incidence.

Next, we probe the impact of spatial separation on the diversity dynamics. Spatial structure is implemented by augmenting the model with a metapopulation element, see Materials and Methods for details. We find that the main effect of space is to protract transitions, such that the transient multimodality of the Hamming distribution lasts longer. The duration of coexistence of strains with different fitnesses is observed to be determined by the transmission rate *β*_*ij*_ between different populations (i.e. with *i* ≠ *j*). In Figure 6–Figure supplement 1, we probe three situations where (relative) inter-population transmission rates are either 0, 10^−4^ or 10^−3^ (with intra-population transmission rates *T*_*ii*_ ≈ 1). We find that with very low (< 10^−3^) transmission rates between populations, spatial structure leads to drawn-out transitions, but that this effect disappears as soon as significant transmission between populations occur. This intuitively makes sense, since the within-population transmission rate will dominate as soon as even a few cases of a new variant have spilled into a population.

It is worth noting that spatial structure in itself cannot lead to the saltational signature seen in Figure 1. By this we mean that a pathogen which evolves gradually (i.e. obeys the weak mutation limit) in multiple spatial patches will not lead to a sudden spike in the Hamming distance once spillover happens. The reason for this is that two spatially remote lineages diverge from each other (in terms of Hamming distance) at approximately the same rate as less geographically distant pairs of lineages.

## Discussion

The pattern of evolution observed in SARS-CoV-2 suggests that transmissibility of the pathogen has mainly increased due to large evolutionary ‘jumps’, rather than due to gradual evolution, something that may turn out to be a signature feature of the pathogen. Our model simulations highlight how this preference for adaptation by saltation may be explained by an ability to overcome epistatic ‘fitness valleys’. The implications for public health are clear; any situation which facilitates such jumps should be treated with heightened awareness. They represent a high risk for the emergence of new, concerning variants which could not have emerged through gradual evolution.

While much attention has been given to the role of immunocompromised individuals, for good reasons, it is important to realize that other probable mechanisms of saltation exist. For instance, consider reverse zoonosis – the transmission from humans to animals. The epistatic landscape may be very different in animals, affording a way of bypassing what would otherwise be troughs in the human SARS-CoV-2 fitness landscape. Reintroduction of the mutated lineage into the human population would then constitute a ‘jump’ in terms of Hamming distance, and potentially also phenotypically. An example of such back-and-forth transmission between human and animal hosts leading to a large number of novel mutations was the so-called Cluster 5 variant, which evolved in mink (*Neovison vison*) in Denmark and subsequently spread to humans (***Hammer et al., 2021***). This mink-derived variant, which was only one of several which escaped into the human population, exhibited 35 substitutions and four deletions in the spike protein alone (***Larsen et al., 2021***).

From a public health perspective, these possible mechanisms have one important thing in common; they underscore the importance of widespread and equitable distribution of up-to-date vaccines, since saltational evolution in disadvantaged or remote populations carries a risk of emergence of new, highly transmissible variants.

The plurality of potential etiologies highlights the need for comprehensive research into the mechanisms which may underlie the observed saltational evolution. Such studies would be most welcome and would have to consider multiple scales, from molecular mechanisms and within-host evolution to the epidemiological dynamics which may contribute to saltations.

The type of analysis performed in this study requires large amounts of sequence data, beyond what could usually be obtained for infectious diseases prior to COVID-19. As shown in Figure 1–Figure supplement 2, a similarly clear and detailed distribution of nucleotide distances could not be obtained for influenza H1N1 or H3N2. This is just one example of how incredibly useful the high level of genomic surveillance achieved for SARS-CoV-2 is, and more generally highlights the potential that extensive sequencing of pathogens holds for advancing phylodynamic understanding across pathogens (***Grenfell et al., 2004***). While many countries have since scaled down the level of testing and sequencing of SARS-CoV-2, scientific insights based on this data will no doubt continue to emerge and have a lasting impact on our understanding of pandemics more broadly.

In our simulations, we have not explicitly modelled any effects of immune memory. We have allowed for new variants with higher effective reproduction numbers to arise, but have not distinguished between whether that advantage stemmed purely from higher infectiousness or from some degree of immune escape. However, it is worth noting that the empirical pattern of punctuated evolution held for every major transition (Figure 1), including the transition to the Alpha variant. When the Alpha variant became dominant, only about 3 infections per 100 people had been recorded in the United Kingdom (***Ritchie et al., 2022***) and vaccinations had not yet begun in earnest. While this is surely an undercount, a general depletion of susceptibles was not a main driver for the success or emergence of the Alpha variant. As such, the punctuated evolutionary pattern does not seem to be hinged on a connection between Hamming distance and evasion of immunity. In the case of the transition to Omicron, immune escape certainly played a role (***Meng et al., 2022***; ***Zhang et al., 2022***), but it would seem that the mechanism of punctuated evolution is more general than that. In ***Meijers et al***. (***2022***), the authors explicitly decompose fitness advantages into intrinsic and antigenic. Introducing a similar distinction in a genotypic fitness landscape model with saltation would be an interesting extension of the present work.

Even in the absence of explicit modeling of immune memory and partial cross-immunity, there are good reasons to believe that saltations will continue to play a role in facilitating the emergence of new variants. As described by ***Plotkin et al***. (***2002***), accumulating immunity changes the fitness landscape of a pathogen over time, lowering some fitness peaks while rendering other peaks relatively more advantageous for the virus. Saltations can then enable the pathogen to reach those fitness peaks. Indeed, it is plausible that high levels of (more or less strain-specific) immunity in a population may increase the rate at which new strains emerge by saltation. Such a connection would further underscore the importance of broadly effective and widely available vaccines as well as any measures which decrease the likelihood of accelerated evolution within hosts, with its risk of seeding saltation events.

As a consequence of the parsimony of our model, we have not explicitly modelled recombination events, but rather assume that each multi-site jump involves a random set of sites. Recombination has been reported in SARS-CoV-2, including – but not limited to – in conjunction with treatment of immunosuppressed patients (***Burel et al., 2022***; ***Duerr et al., 2022***; ***Varabyou et al., 2021***; ***Focosi and Maggi, 2022***). Future work could explore the implications of allowing for recombination events in this type of model.

The influenza model of ***Koelle et al***. (***2006***), which gives rise to Hamming dynamics reminiscent of the saltation-free simulations of Figure 4, does so in a very different way. There, it is assumed that the pathogen explores a *neutral network* (a set of antigenically and fitness-wise equivalent genotypes which are connected by one-mutation neighbours (***Newman and Engelhardt, 1998***)) in the vicinity the prevailing strain. This goes on until the ‘random walk’ happens upon a configuration which is substantially antigenically different from the prevailing cluster, albeit connected to it by a single mutation. Once this happens, a new cluster emerges which has only limited cross-immunity with the prevailing strain. Since all steps along the way are small, the new variant will be very close (genotypically) to a member of the previous cluster. Consequently, this type of dynamics does not produce abrupt spikes in Hamming distance, such as the ones shown in Figures 1 and 3.

There are a few models in the literature that seek to address the connection between saltation, epistasis and the likelihood of emergence of new variants (***Katsnelson et al***. (***2019***) and ***Smith and Ashby*** (***2022***), the latter of which is based on the model by ***Gog and Grenfell*** (***2002***)). However, in contrast to existing theoretical studies, we address the empirical temporal development of diversity and propose a model which can directly replicate the main features of that distribution.

We have focused on capturing the main features of the evolution of SARS-CoV-2 as parsimoniously as possible and although we have explored a number of biologically motivated extensions, our model still represents a theoretical foundation upon which more sophisticated models can be built. There is much to be done in terms of understanding and modelling the precise fitness landscape of SARS-CoV-2, including its dependence on host immunity history. More broadly, an increase in genomic surveillance across multiple pathogens will doubtlessly lead to new insights into the diversity dynamics of other pathogens. This would not only enable research into the evolution of individual pathogens, but allow us to question how co-circulating pathogens affect the diversity dynamics of one another.

## Methods and Materials

### Temporally resolved Hamming distributions from sequence data

In this section, we describe the data processing workflow which was used to generate the Hamming distance plots of Figures 1 and 2. We have used the open, GenBank-derived dataset of aligned SARS-CoV-2 sequences from Nextstrain (***Hadfield et al., 2018***). In our main analysis, we have used UK sequences from the 1st of March 2020 onwards. See also Figure 1–Figure supplement 1 for an illustration of the following workflow:

- For each day *t* in the interval:
  - Select 5,000 random pairs of whole-genome sequences (i.e. 10,000 sequences) obtained within a 1-week time window starting on day *t*.
  - For each pair of sequences *s*_*i*_ and *s*_*j*_:
    * Go through both sequences, site by site, and record the number of differences between them, *H*_*ij*_. This is the pairwise Hamming distance.
  - Compute a probability density/histogram *p*_*t*_(*H*) based on the observed Hamming distances {*H*_*ij*_}.

It is then this function, *p*_*t*_(*H*), that it plotted in Figure 1A. In practice, we have used the metadata provided by Nexstrain, which contains fields describing the nucleotide differences relative to the reference strain Wuhan-Hu-1 (GenBank reference sequence accession number MN908947.3), rather than operating directly on the whole-genome sequences. Numerically, this makes no difference, but it affords a large increase in performance, since it allows us to avoid processing unchanged regions of the genome, which do not contribute to the Hamming distance.

For Appendix 2 Figure 1, which instead shows the distance to the reference sequence (the ’absolute’ Hamming distance), the above workflow is slightly altered:

- For each day *t* in the interval:
  - Select 5,000 random whole-genome sequences obtained within a 1-week time window starting on day *t*.
  - For sequences *s*_*i*_:
    * Go through the sequence, site by site, and record the number of differences *H*_*i*_ between *s*_*i*_ and the reference sequence. This is the absolute Hamming distance.
  - Compute a probability density/histogram *p*_0,*t*_(*H*) based on the observed Hamming distances {*H*_*i*_}.

It is then this function, *p*_0,*t*_(*H*), that it plotted in Appendix 2 Figure 1.

### Branching model with saltational evolution

The mechanistic model developed for this study is a discrete-time branching model coupled to a genotypic fitness landscape model.

In the simulations of Figures 3 and 4, we assumed a constant prevalence, for simplicity. This amounts to keeping the mean effective reproductive number across the population at unity. In Figure 6 we relax this assumption and explore a version of the model with epidemic dynamics. We start by documenting the constant-prevalence version of the model, as well as the genotypic fitness landscape element, before we go on to describe how we incorporate SIRS dynamics and spatial structure.

### Evolutionary branching model with constant prevalence

In the model, each new generation of infections consists of a fixed number of individuals, *N*, and generations do not overlap. Consequently, there are *N* infected individuals at any given time. Each infected individual *i* has an associated bit-string *G*_*i*_ of length *L*, representing the genome of the pathogen. We do not explicitly model any within-host diversity, as we are only interested in the genome of the pathogen that is eventually transferred during transmission.

At each time step (corresponding to one generation), a new random individual *i* is repeatedly selected and allowed to infect a number *z*_*i*_ of new individuals, selected from a Poisson distribution with mean *R*_*i*_, i.e. *z*_*i*_ ∼ *Pois*(*R*_*i*_). This continues until a total of *N* new transmissions have occurred in that generation, ensuring that the prevalence is kept constant. At transmission, the pathogen genome of the infector is copied to the infectee. The personal reproductive number *R*_*i*_ is deter-mined by the fitness of the bit-string *G*_*i*_, the details of which are discussed in the next subsection. In each newly infected individual, there is a risk of mutation. The number of point mutations *m*_*i*_ that occur within the *i*’th host is drawn from a distribution. In the case of homogeneous mutation (i.e. absence of saltation), *m*_*i*_ is drawn from a Poisson distribution characterized by a mutation rate *μ*_0_ < 1. Saltation, on the other hand, is simulated by drawing *m*_*i*_ from a bimodal distribution characterized by two different mutation rates/sizes *μ*_0_ and *μ*_1_, ensuring that an outsized amount of mutation can take place within a single host on rare occasions. Concretely, we have used the distribution *P*_*s*_(*m*) given by:

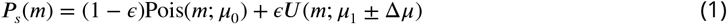

Where *U* (*m*; *μ*_1_ ± Δ*μ*]) is the uniform distribution centered on *μ*_1_ with half-width Δ*μ*, and Pois(*m*; *μ*_0_) is a Poisson distribution with mean *μ*_0_. *ϵ* ≪ 1 is a small dimensionless quantity measuring the frequency of saltational mutation. The parameter *μ*_0_ gives the rate of non-saltational mutation while *μ*_1_ is the typical size of a saltation.

We use this simple bimodal distribution out of convenience, but our results do not change qualitative if another bimodal distribution is used.

Once the quantity *m*_*i*_ has been drawn, a number *m*_*i*_ of random bit flips are then performed in the genomic bitstring *G*_*i*_, each flip corresponding to a point mutation.

### Modelling sign epistasis

Before simulations start, a number of *N*_*e*_ of ‘epitopes’ (regions in the genome on which fitness depends), each of length *L*_*e*_, are designated. We assume non-overlapping epitopal regions and thus require *L*_*e*_*N*_*e*_ ≤ *L*.

Within each epitope, a number *N*_*H*_ of highly fit combinations are assigned. We have assumed *N*_*H*_ = 1 for all of our simulations, but since the general *N*_*H*_ case is no more complicated, we include the parameter here. The fitness of each combination is measured in terms of its contribution *δR*_*H*_ to the individual reproductive number. In general, *δR*_*H*_ for each combination may be drawn from a distribution *P*_*H*_ (*δR*_*H*_) to allow for a variety of combinations with different fitness values.

Tunable sign epistasis is modeled by assigning a fitness contribution *δR*_*L*_ ≤ 0 to each combination which lies within a Hamming distance *d*_0_ of a high-fitness combination. The overall fitness of a given genotype is then obtained by adding up the contributions for each of the *N*_*e*_ epitopes:

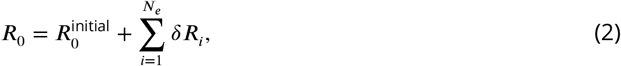

with the constraint that *R*_0_ ≥ 0. In practice this constraint is enforced by letting

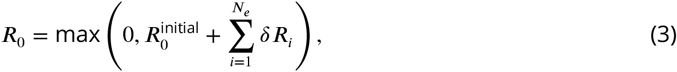

High sign epistasis is then achieved when *d*_0_ > 1 and *δR*_*L*_ ≪ 0. However, the model also allows for incomplete or partial sign epistasis: if *δR*_*L*_ for each combination is drawn from a distribution *P*_*L*_(*δR*_*L*_) which has support at *δR*_*L*_ = 0, then each peak in the fitness landscape will not be completely surrounded by troughs. In other words, in that case it may be possible to evolve to a highly fit variant through a series of single point mutations without suffering decreased fitness in the process.

Unless otherwise specified, we run our simulations with the parameter values given in Table 1

**Table 1.** Model parameters and their values.

| Parameter | Description | Value<br>(base case) |
| --- | --- | --- |
| $L$ | Genome length (bits) | 1000 |
| $L_e$ | Length of each epitopal sequence | 5 |
| $N_H$ | Number of highly fit configurations of each epitope | 1 |
| $N_e$ | Number of epitopes | 5 |
| $d_0$ | Hamming distance within which genotype is deleterious | 3 |
| $\langle \delta R_H \rangle$ | Avg. fitness advantage (change in $R$) due to beneficial genotype | 1 |
| $\langle \delta R_L \rangle$ | Avg. fitness contribution (change in $R$) due to deleterious genotype | -1 |
| $\mu_0$ | Base mutation rate (for whole genome) | 0.3 |
| $\epsilon$ | Frequency of saltation | 0 or 0.0001 |
| $\mu_1$ | Typical size of saltations | 150 |
| $\Delta\mu$ | Half-width of saltation size distribution | 50 |
| $T_{ij}$ | Relative transmission rate betw. populations $i, j$ Subject to $\sum_j T_{ij} = 1$. | 0-1 |
| $\omega$ | Rate of waning of immunity in SIRS simulations | 0.1 |

### Incorporating SIRS dynamics

In the simulations of Figures 3 and 4 we assumed constant incidence, meaning that the number of infected within any given generation was *I*(*t*) = *I*_0_ with *I*_0_ a constant (thus, prevalence was constant as well). However, to relax this assumption we incorporate susceptible-infected-recovered-susceptible (SIRS) dynamics.

We continue to model the infected individuals by a branching process, but now also keep track of the number of susceptible (S(*t*)) and recovered (*R*(*t*)) individuals in each generation *t* of the outbreak.

The simulations proceed as follows. At time *t* = 0, let:

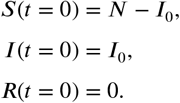

Then, at each subsequent time step *t*, the transmission probability is modulated by a factor S(*t*)/*N* representing susceptible depletion. Whenever transmission does occur, the number of susceptibles decrease by one while the number of infected individuals increase correspondingly. When an individual is recovered (i.e. after one generation of being infected), *I*(*t*) is decreased by one while *R*(*t*) increases by one.

Each recovered person has a constant probability rate *ω* for becoming susceptible once again. In other words, this is modeled as a Poisson process with rate *ω*. Note that this not only corresponds to waning of immunity, but also to any other mechanism by which a recovered individual may become replaced by a susceptible one (such as population turnover). However, we will refer to *ω* as the rate of waning. In our simulations (Figure 6 and Figure 6–Figure supplement 1), we set 1/*ω* = 10 meaning that duration of immunity averages 10 generations. This figure is not supposed to reflect any particular value for SARS-CoV-2, but is rather used to illustrate the robustness of the pattern of punctuated evolution to waning immunity. In the interest of simplicity, we have ignored any seasonal effects on transmission. We consider this a reasonable simplification, both due to the conceptual nature of our model and the understanding that susceptible dynamics rather than seasonality is the major limiting factor in the pandemic phase (***Baker et al., 2020***).

### Spatial structure

We implement a minimal model of spatial structure by incorporating a metapopulation element. Let there be *n*_pops_ populations, each with total population *N*_*i*_ (*i* ∈ {1, …, *n*_pops_}). At time *t* = 0, let the number of susceptible, infected and recovered individuals in each population be given by:

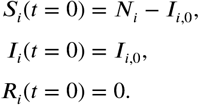

In our simulations (Figure 6–Figure supplement 1) we assume identical population sizes, *N*_*i*_ = *N*/*n*_pops_, and an initial equipartitioning of infected individuals *I*_*i*,0_ = *I*_*i*_/*n*_pops_, where Σ_*i*_ *N*_*i*_ and *I*_0_ = Σ_*i*_ *I*_*i*,0_.

The transmission rate between populations is then determined by the matrix elements *β*_*ij*_ = *βT*_*ij*_ where each element *T*_*ij*_ gives the relative transmission rate from population *i* to *j* and *β* represents the transmissibility of the strain the infected individual carries. We assume that **T** is a symmetric matrix, *T*_*ij*_ = *T*_*ji*_.

In Figure 6–Figure supplement 1 we took **T** to have the following form:

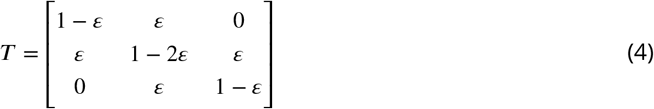

with ε at either 0 (panel A), 10^−4^ (panel B) or 10^−3^ (panel C). This corresponds to a linear layout of three populations, with transmission occurring only between adjacent compartments.

### Modelling decreasing absolute Hamming distance

As described in Appendix 2, the typical Hamming distance between circulating genomes and the ancestral variant is not necessarily monotonically increasing with time. We call this distance the *absolute* Hamming distance, in contrast to the pairwise distance between concurrently circulating genomes which we call the *relative* Hamming distance (to reflect that the absolute Hamming distance is measured with respect to a fixed point in the genomic space).

We begin by describing a very simple variation upon the model which has the effect of allowing the absolute Hamming distance to decrease (as well as increase) at variant transitions. In this section, we assume a constant prevalence of *N* infected individuals.

Assume that a fraction *p*_*d*_ of the first generation (i.e. *p*_*d*_ *N* individuals) have prolonged infections, lasting *τ*_*d*_ typical generations before onward transmission. Assume furthermore that mutations happen at a rate *μ*_*d*_ for these individuals, such that a number of point mutations, *μ*_*d*_ *τ*_*d*_, occurs before onward transmission. Here, *μ*_*d*_ is the mutation rate associated with these prolonged infections. The rest of the population is assumed to be homogeneous with respect to the occurrence of mutations, all possessing a mutation rate *μ*_0_. We draw *τ*_*d*_ from a uniform distribution with support throughout the entire simulation (which is assumed to have duration *t*_*f*_), *τ*_*d*_ ∼ *U* (*τ*_*d*_ ; *t*_*f*_ /2 ± *t*_*f*_ /2). Furthermore, the fitness advantage *δR*_*H*_ of different epitope configurations was drawn from a uniform distribution as well, to avoid fitness degeneracy (multiple equally fit variants).

This simple modification of the model enables a non-monotonic time development of the absolute Hamming distance, as shown in Appendix 2 Figure 2B, while preserving the dynamics of relative Hamming distance shown in Figure 3.

This is of course a highly simplistic variation upon the base model, but it serves to show that pro-longed infection or introduction of (mutated versions of) previous variants can account for absolute Hamming distance sometimes decreasing at variant transitions.

## Data Availability

All data and code produced in the present study are available upon reasonable request to the authors.

## Acknowledgments

We would like to thank the members of the Grenfell and Levin Labs at The Department of Ecology and Evolutionary Biology, Princeton University, for fertile plenary discussions. We would also like to thank Arne Traulsen and Chadi M. Saad-Roy for enlightening discussions pertaining to the formulation of our model and Christian Berrig and Viggo Andreasen at the PandemiX Center, Roskilde University, for much appreciated comments on data visualization.

## Research Funding

BFN and LS received funding from the Carlsberg Foundation under its Semper Ardens programme (grant #CF20-0046, BFN and LS). BTG received funding from the Flu Lab. SAL acknowledges the support of the the C3.ai Digital Transformation Institute and Microsoft Corporation, Gift from Google and the National Science Foundation (CNS-2027908, CCF1917819).

## Appendix 1

### Diversity dynamics based on US sequences

In the main text, we based our analysis on sequence data from the United Kingdom, since the UK genomic surveillance was extensive. However, the overall results remain the same if one instead considers United States sequence data. The exact timing of the variant transitions differ – most notably, the interval between the Alpha and Delta transitions is shorter – but the qualitative features are similar. Variant transitions continue to be associated with large jumps in Hamming distance, indicative of saltation. The temporal Hamming distribution for the US is shown in Figure 1. Due to the shorter interval, the Alpha and Delta transitions are not as clearly separated as in the UK data, and the Alpha transition appears somewhat drawn out. As demonstrated in Figure 6–Figure supplement 1, this could be a result of the presence of greater spatial effects.

**Appendix 1 Figure 1.**
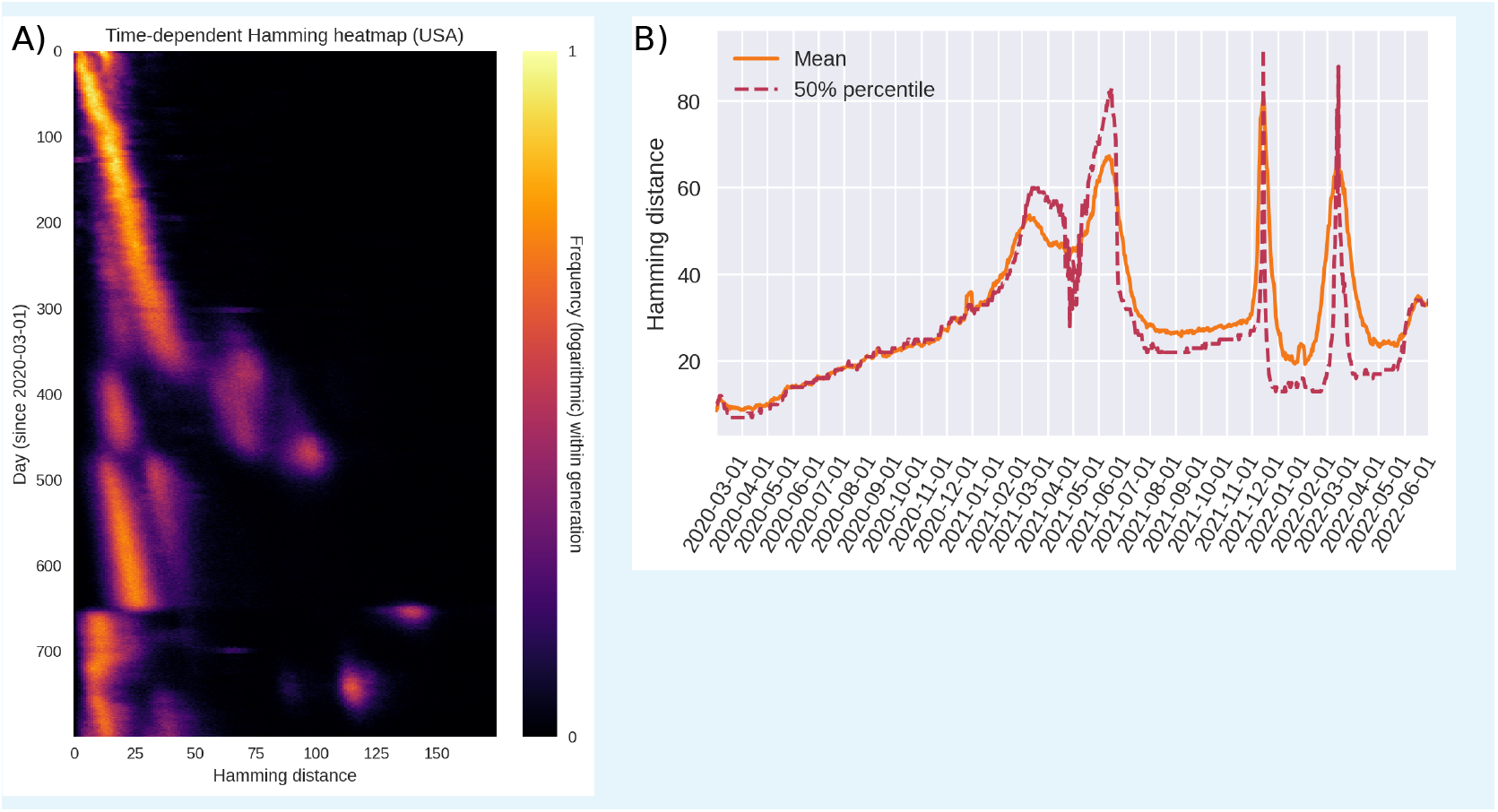
Hamming distance analysis analogous to that of Figure 1, except based on United States sequence data, rather than British sequences. **A)** The full, time-dependent Hamming distribution (US data, GenBank via Nextstrain). **B)** Time evolution of the mean and median pairwise Hamming distances.

## Appendix 2

### Fitting the origin-centered Hamming distribution

Our analysis is mainly centered around the dynamics of diversity, as measured by the dissimilarity of SARS-CoV-2 genomes which are in circulation at a given point in time (Figure 1). Our primary goal was to formulate a dynamical model which could qualitatively replicate this pattern as parsimoniously as possible.

In this section, we additionally consider the distance between circulating genomes and the *origin* (meaning the sequence Wuhan-Hu-1, GenBank reference sequence accession number MN908947.3). In Appendix 2 Figure 1, we show the distribution of distances between circulating genomes and this reference sequence, which we will call the *absolute* Hamming distance. The data analysis workflow in creating this plot is analogous to that detailed in Figure 1–Figure supplement 1, but instead of continuously picking *pairs* of genomes, we pick single genomes which are then compared with the reference genome. This also means that, given *N* genomes within a given time window, there can be only *N* data points with this method. This is in contrast to the pairwise comparison in Figure 1, where *N* genomes give rise to *N*(*N* − 1) ∼ *N*^2^ possible pairings.

In Appendix 2 Figure 1, we see that the absolute Hamming distance at first increases approximately linearly with time until the Alpha transition, at which point a large jump in the absolute Hamming distance is observed. Interestingly, the transition from Alpha to Delta is associated with a slight decrease in this distance. The Omicron transition is again associated with a large increase in the absolute Hamming distance.

**Appendix 2 Figure 1.**
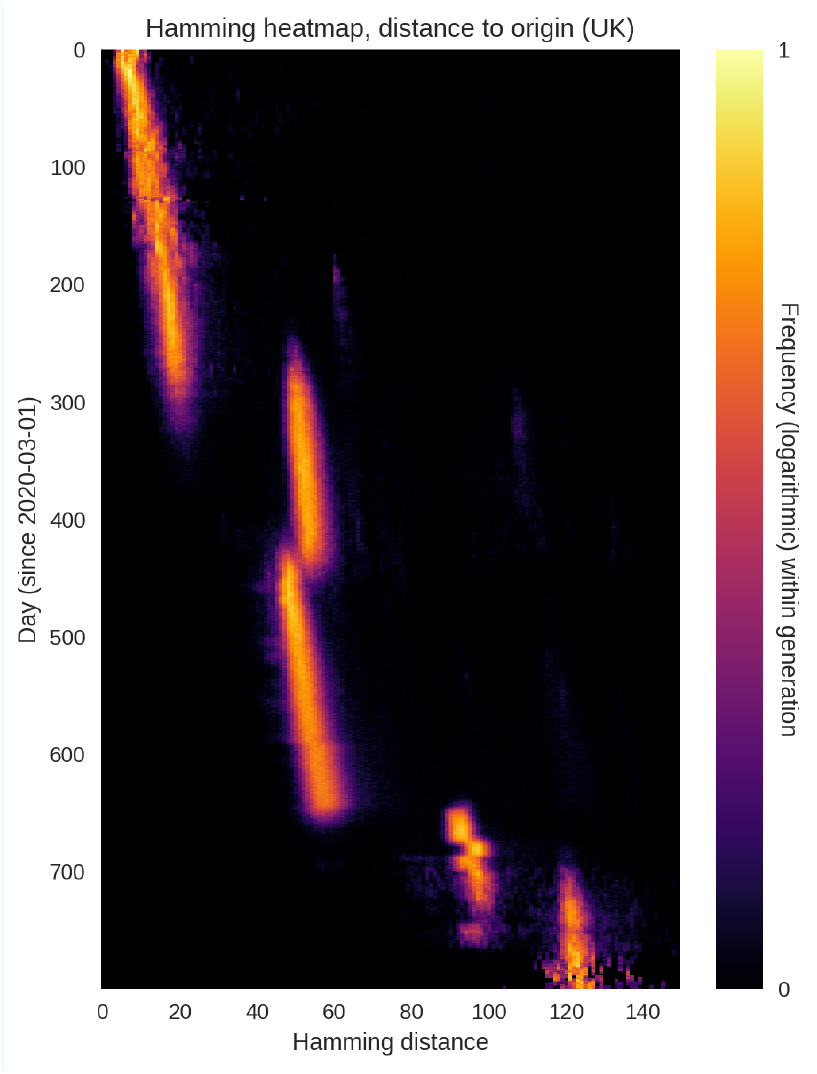
**Time evolution of the *absolute* Hamming distance**, i.e. the distance to the origin (defined as the Wuhan-Hu-1 reference sequence). UK sequence data.

One possible explanation for such a decrease in absolute Hamming distance is that pro-longed infections with an earlier variant have occurred in some individuals or a population, albeit without accompanying accelerated evolution. Once the unobserved lineage spills into the sampled population, it may lead to a variant transition despite being closer to the ancestral variant than to the currently dominating one.

In terms of our model, this kind of dynamics can be quite simply incorporated. While it is not impossible to see decreases in absolute Hamming distance in the simple model formulation behind Figure 3, it is highly statistically unlikely, since each saltation happens on the basis of the genomes present in the previous generation.

The very simplest way to incorporate the possibility of decreasing absolute Hamming distance is to assume that a certain fraction of the population are initially infected with the ancestral strain, but that their infections are prolonged (and that they thus only transmit the disease much later). If the pathogen undergoes mutation within these hosts, it is possible to obtain a pattern such as the one shown in Appendix 2 Figure 2B. As evidenced by panel A of that same figure, the pairwise ’relative’ Hamming distance between genomes in the same generation is not qualitatively affected by this addition to the model. Transitions are still driven by large saltations, and the (relative) Hamming distance is characterized by periods of linear growth punctuated by large increases and subsequent collapses in diversity.

The technical details of this addition to the model can be found in the Materials and Methods section.

While allowing for persistent infections with the initial variant is of course a very simple variation on the base model, it does show that prolonged infections with previous variants can account for occasional sudden decreases in the absolute Hamming distance.

**Appendix 2 Figure 2.**
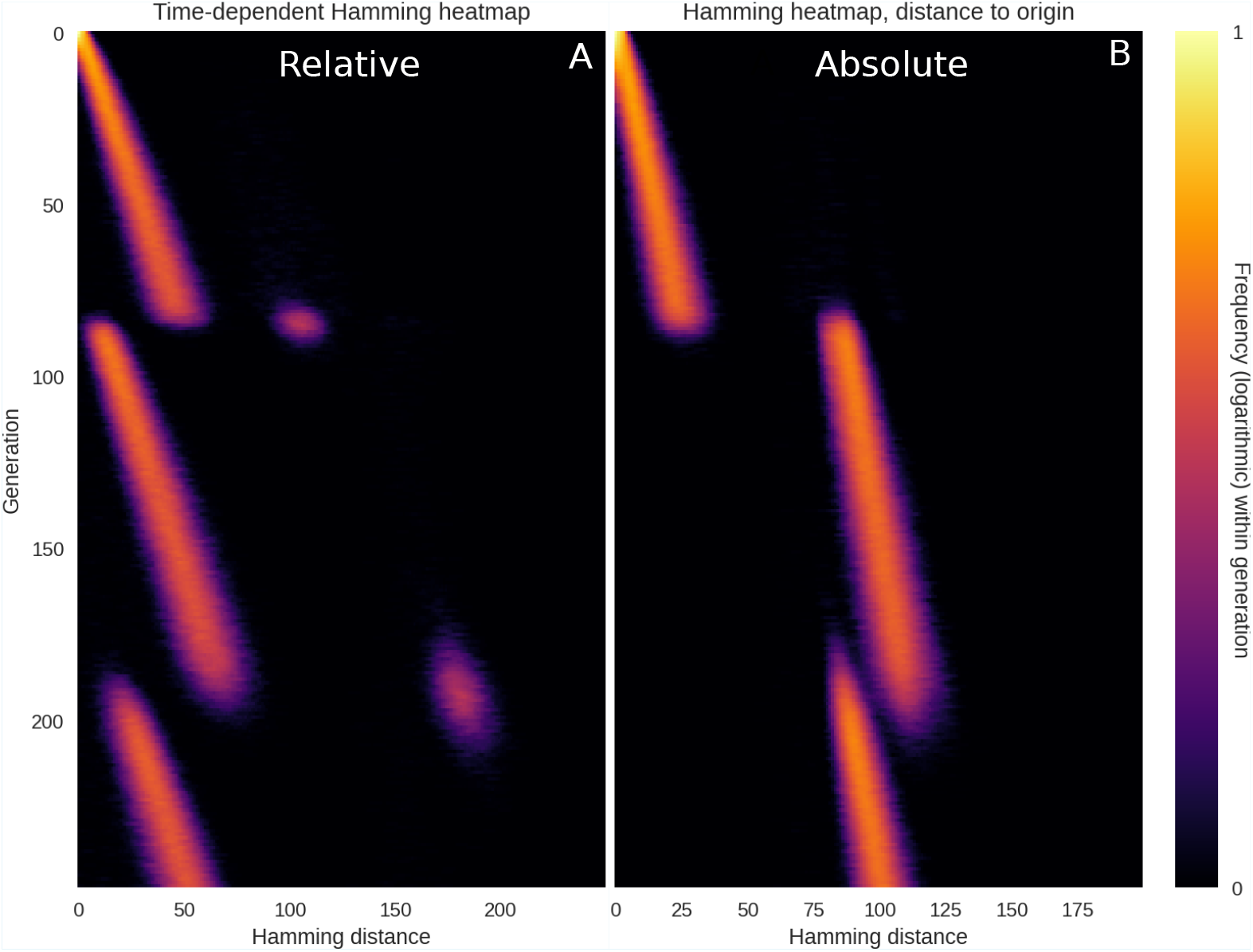
Simulation with persistent infections with original variant. When the original variant persists in a small fraction of the population, absolute Hamming distance can decrease at variant transitions. **A)** The ‘relative’ Hamming distance, the same quantity that was plotted in e.g. main text figures 1 and 3. It measures the dissimilarity between concurrently circulating pathogen genomes. **B)** The ‘absolute’ Hamming distribution, measuring the distance between circulating pathogen genomes and the reference sequence, namely that of the initial variant.

---

**Figure 1–Figure supplement 1.**
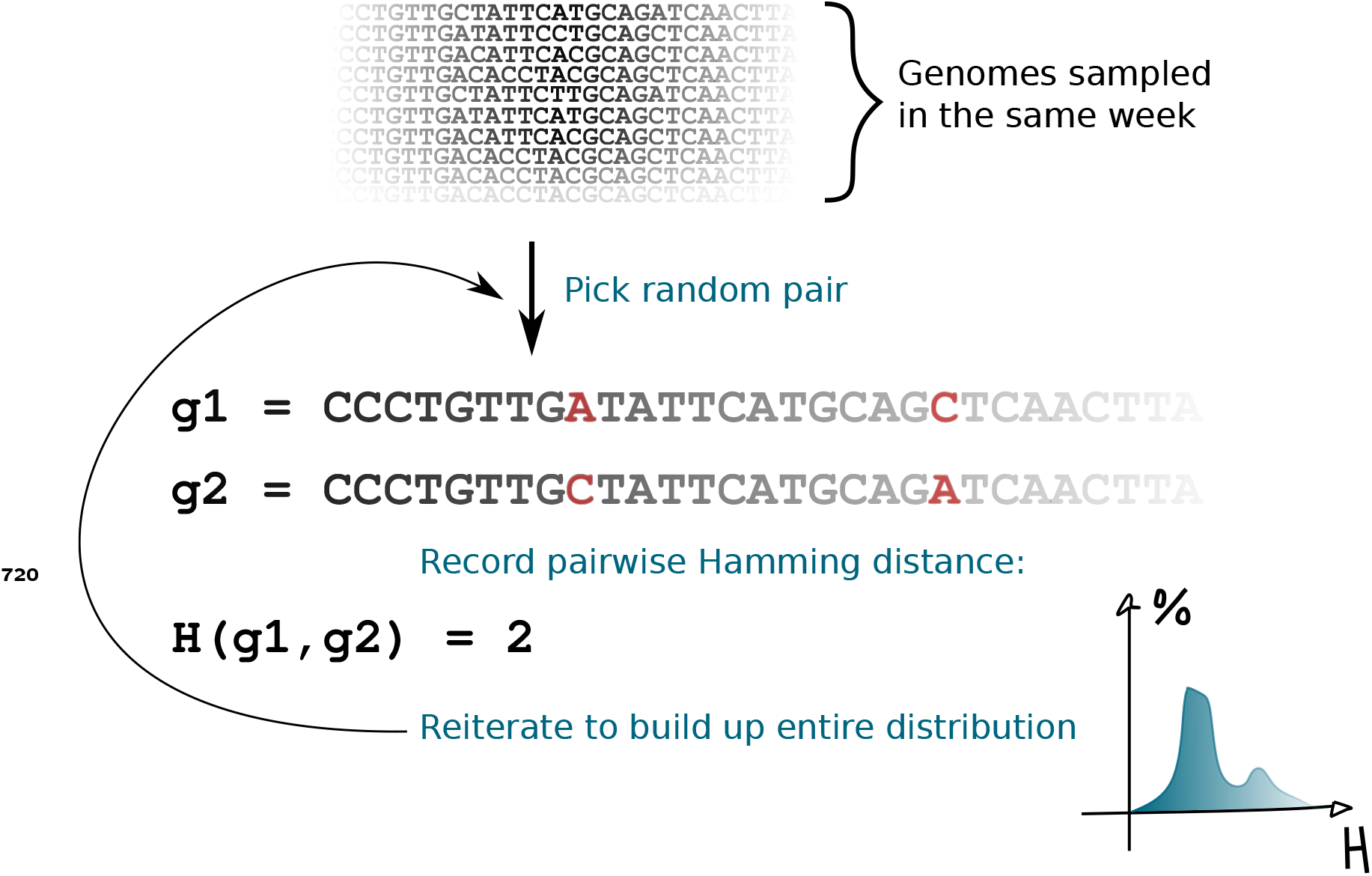
Data analysis workflow. To generate the Hamming distribution for a given point in time, all sequences sampled within a week-long window starting on the given day are pooled. Then, pairs of sequences are repeatedly selected at random from this sequence pool, and the pairwise Hamming distance (number of sites which differ) is computed. All the computed Hamming distances are then pooled and a distribution (histogram) is generated.

**Figure 1–Figure supplement 2.**
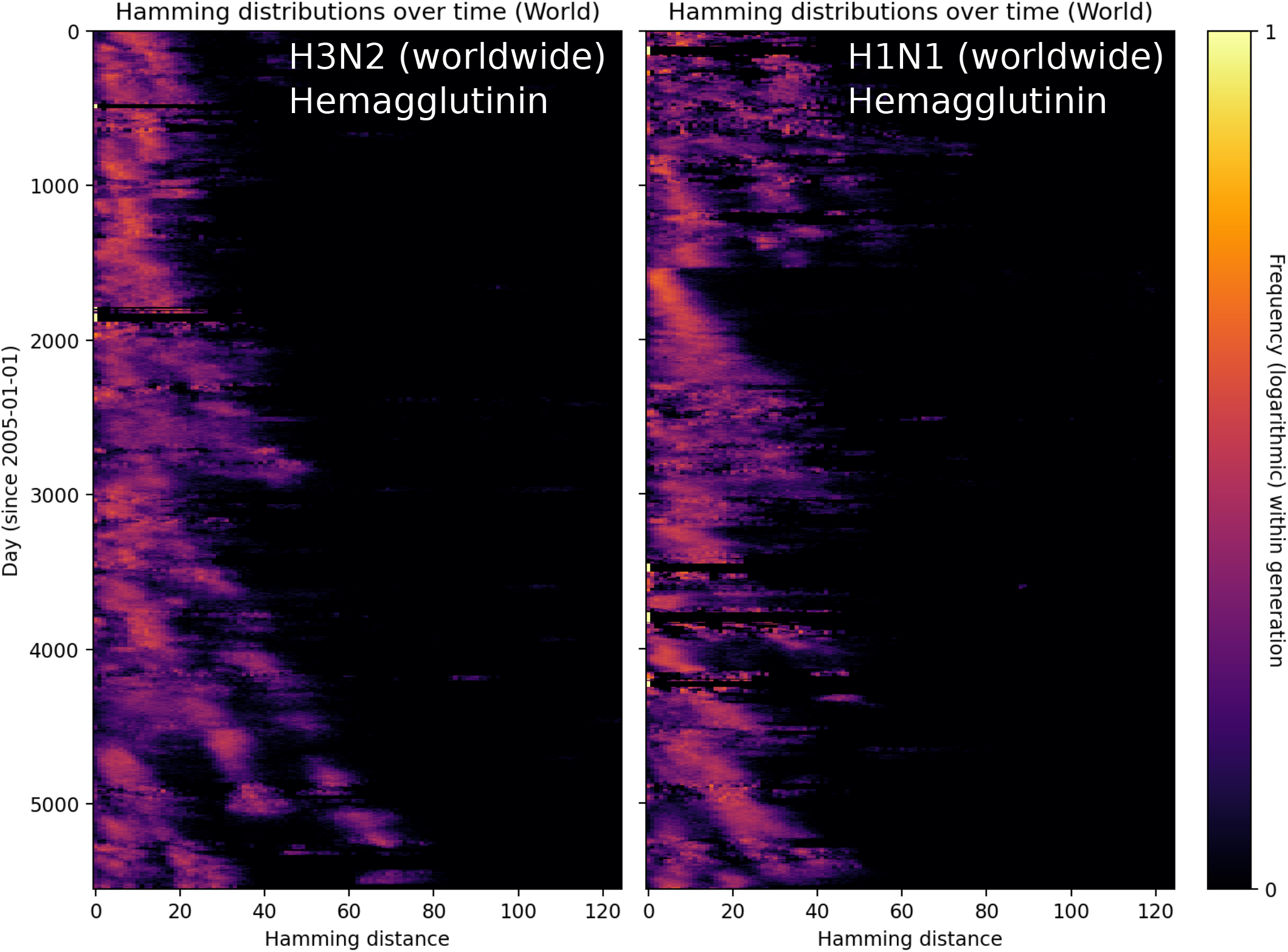
Hamming distributions for influenza H3N2 and H1N1. With influenza, the amount of genomic surveillance data is much more limited and the temporal Hamming distributions are much less well-defined. In order to have enough data for each time point, a sampling window of 30 days was used here, as opposed to the 7 days used for SARS-CoV-2 in the main text.

**Figure 3–Figure supplement 1.**
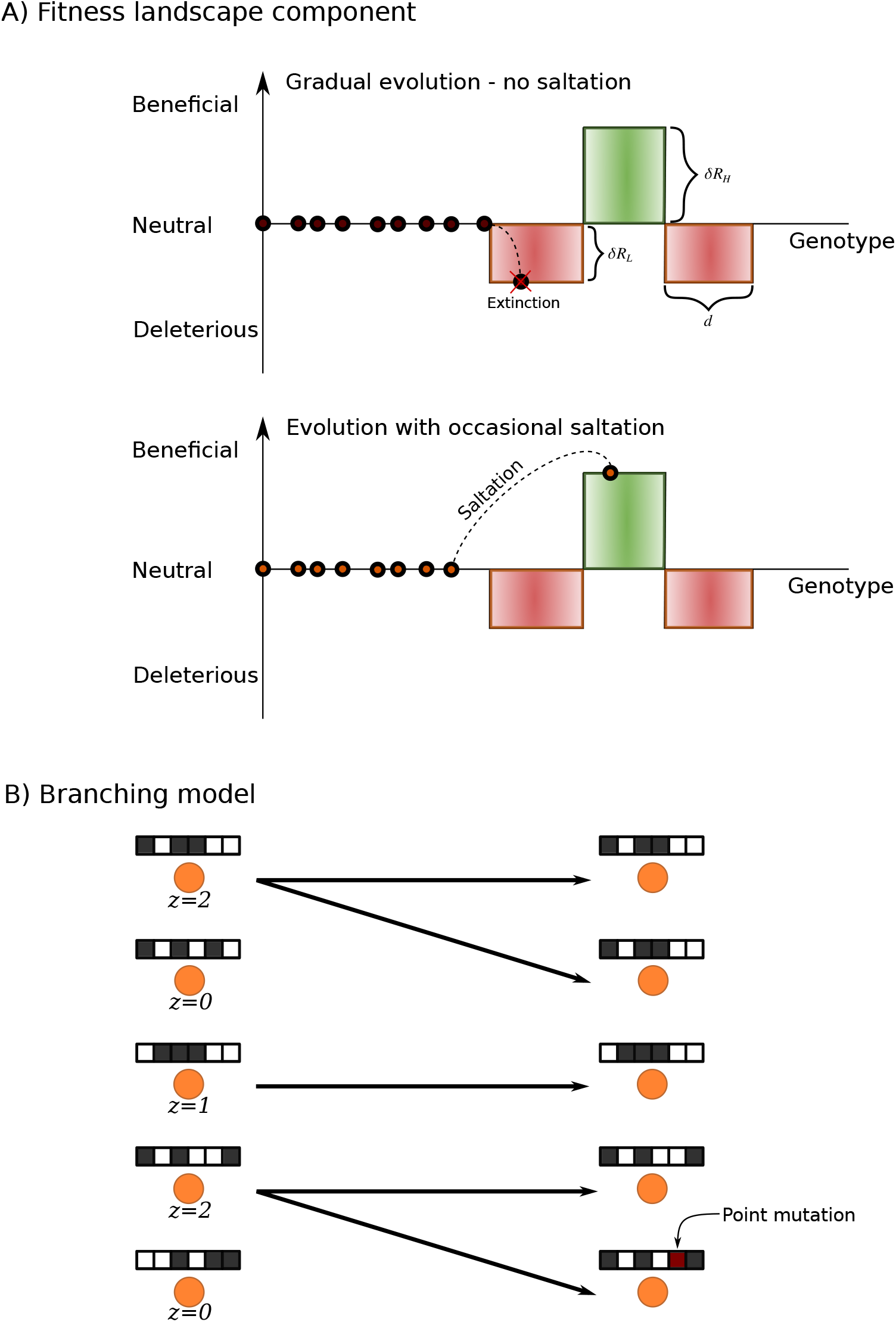
Model schematics. **A)** The fitness landscape and epistasis components of the model. The majority of the fitness landscape is assumed neutral. In the case of gradual evolution devoid of saltation (top), the pathogen performs a random walk in this neutral space until it hits upon a deleterious configuration. As a model of sign epistasis, beneficial configurations are surrounded by deleterious ones. In the case of gradual evolution, the deleterious regions are unlikely to be traversed before the lineage dies out. However, in the case of saltational evolution (bottom), several point mutations may occasionally happen in the same genome within the same generation, leading to a jump which can enable the pathogen to bypass a deleterious region. Note that this is only a 1-dimensional conceptual representation of a highly multidimensional fitness landscape. **B)** In each generation of the branching model, each individual stochastically infects *z* new individuals. Upon infection, the pathogen genome (depicted as a string of black and white squares) is transferred. Occasionally a point mutation will occur, as indicated in the lower right genome. In the case of saltation (see panel A), multiple such point mutations can occur within the same genome in the same generation.

**Figure 3–Figure supplement 2.**
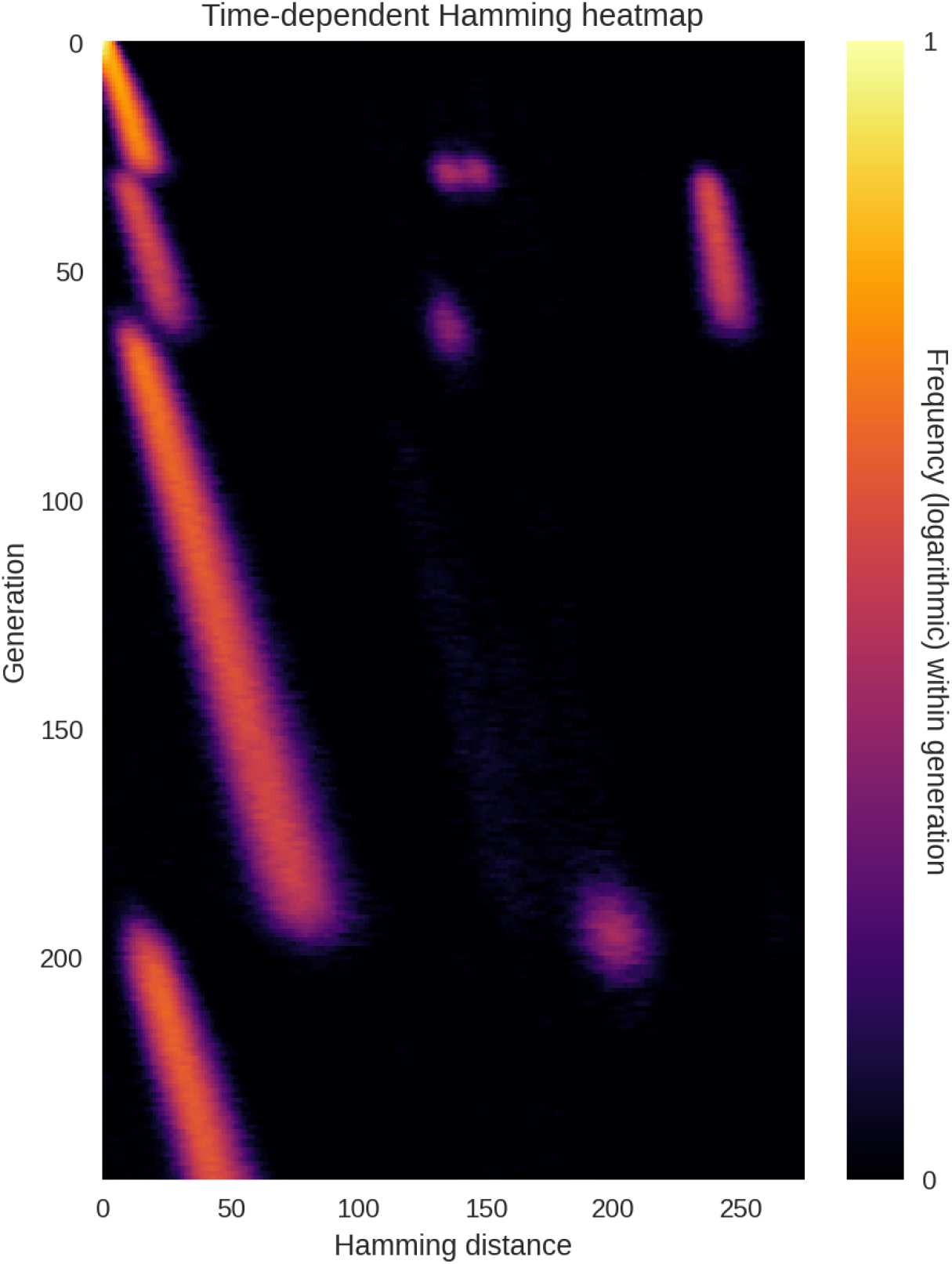
Temporary coexistence of two equally fit variants.

**Figure 4–Figure supplement 1.**
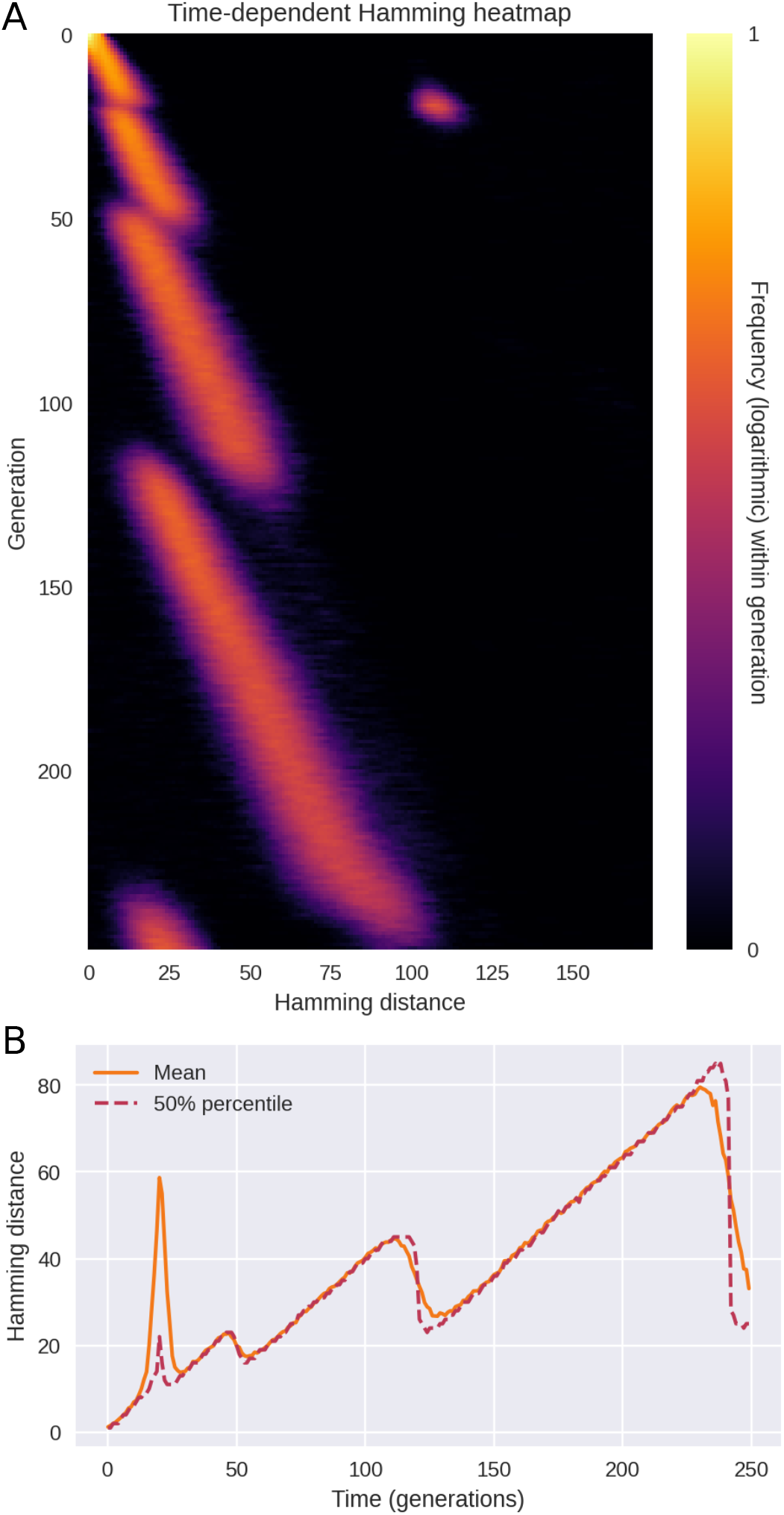
Saltational evolution in the absence of sign epistasis. When saltational evolution is allowed, but epistasis is absent or very weak, a mixture of qualitatively different transitions occur. Some resemble the diversity spikes seen in Figure 3, but more commonly transitions will involve a gradual, linear increase in diversity followed by a collapse, as seen in Figure 4. **A)** Time evolution of the Hamming distance distribution. For each generation indicated on the vertical axis, the colour encodes the histogram of Hamming distances between genomes within that generation. **B)** Time evolution of the mean and median Hamming distance between genomes present in any given generation of the model simulation. In these simulations, *δR*_*L*_ = 0 (no epistasis) while saltations were of typical size *μ*_1_ = 150.

**Figure 6–Figure supplement 1.**
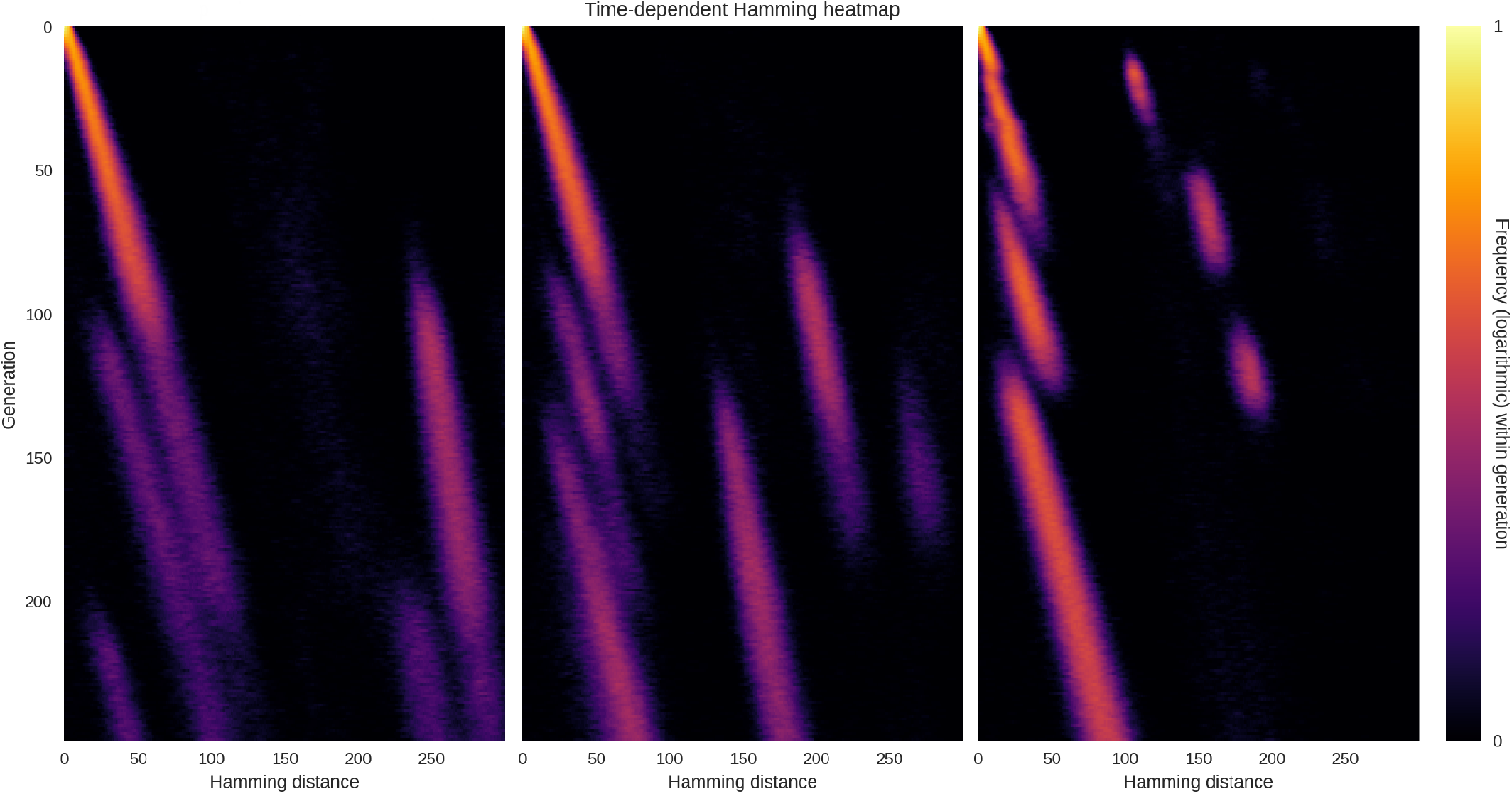
Spatial structure leads to prolonged transition. Here we simulate the same SIRS dynamics as in Figure 6, but in a metapopulation consisting of three subpopulations. The within-population transmission rate *T*_*ii*_ ≈ 1 (*i* ∈ {1, 2, 3}) is assumed much greater than the between-population transmission rate *T*_*ij*_ (with *j* = *i* ± 1). **(A)** With inter-population transmission rate *β*_*i,i*±1_ = 0, mutations never spread from one population to another and coexistence of variants with different fitness can last indefinitely. **(B)** With an inter-population transmission rate of 10^−4^, transitions are severely prolonged but coexistence of variants with different fitness values does not last indefinitely. **(C)** At an inter-population transmission rate of 10^−3^, transitions are only moderately prolonged compared to the non-spatial dynamics of Figure 6.

## Footnotes

1 By ancestral variant, we mean the lineages circulating before the Alpha transition, whether including the D614G substitution or not (***Hou et al., 2020***; ***Isabel et al., 2020***).

2 The BA.2→BA.5 transition was also muddled somewhat by the B.1.1.529 subvariant briefly making up as much as 10% of UK sequences (***Hodcroft, 2021***)

3 Note that time in the model is measured in generations, meaning that constant incidence and prevalence both hold.

